# Common infections and neuroimaging markers of dementia in three UK cohort studies

**DOI:** 10.1101/2023.07.12.23292538

**Authors:** Rebecca E Green, Carole H Sudre, Charlotte Warren-Gash, Julia Butt, Tim Waterboer, the Insight 46 study team, Alun D Hughes, Jonathan M Schott, Marcus Richards, Nish Chaturvedi, Dylan M Williams

## Abstract

**Introduction:** We aimed to investigate associations between common infections and neuroimaging markers of dementia risk (brain volume, hippocampal volume, white matter lesions) across three population-based studies.

**Methods:** We tested associations between serology measures (pathogen serostatus, cumulative burden, continuous antibody responses) and outcomes using linear regression, including adjustments for total intracranial volume and scanner/clinic information (basic model), age, sex, ethnicity, education, socioeconomic position, alcohol, BMI, and smoking (fully adjusted model). Interactions between serology measures and APOE genotype were tested. Findings were meta-analysed across cohorts (N_main_=2632; N_APOE-interaction_=1810).

**Results:** Seropositivity to JC virus associated with smaller brain volumes in basic models (ß=-3.89ml[-5.81,-1.97],p_adjusted_<0.05); these were largely attenuated in fully adjusted models (ß=-1.59ml[-3.55,0.36], p=0.11). No other relationships were robust to multiple testing correction and sensitivity analyses, but several suggestive associations were observed.

**Discussion:** We did not find clear evidence for relationships between common infections and markers of dementia risk. Some suggestive findings warrant testing for replication.

## Introduction

Common infections have been associated with a variety of non-communicable diseases, with a role in dementia suspected ^1^. Despite primary infection typically occurring earlier in the life course, a number of pathogens are able to cause persistent infections; for example through chronic infection or by remaining latent and reactivating at later stages ^2^. As infections may be preventable or treatable, a better understanding of their role in dementia risk could inform our knowledge of high-risk groups and possible priorities for intervention.

While the extent to which common infections contribute to dementia risk is still unclear, multiple pathways have been hypothesised. Neurotropic pathogens – such as herpes simplex virus (HSV) – are able to directly infect cells in the central nervous system, possibly triggering amyloid-ß pathology ^3^, neuroinflammation, and neuronal loss ^4^. Infection or cumulative infection burden could additionally drive systemic changes, including inflammation ^5^, which have been implicated in dementia aetiology ^6, 7^. Nevertheless, previous studies have primarily focussed on a few hypothesis-driven pathogens or pathogen families, and findings have been mixed ^8^. This may be due to challenges in defining previous infection in the absence of large-scale serology data, confounding and, as the prodromal and clinical phases of dementia may affect immune competency, reverse directionality.

Examining relationships between multiple common infections and subclinical markers of dementia prospectively – across well-characterised population-based studies – could facilitate a more comprehensive understanding of their role in dementia risk. In addition, assessing whether pathogen exposure may interact with established risk or protective factors of dementia could provide insights on possible at-risk groups. For example, APOE genotypes – the strongest genetic risk (for APOE e4 carriers) and protective (for APOE e2 carriers) factors for late-onset dementia – are suggested to modify associations of specific infections or burden with dementia risk ^9–11^.

Recently, a validated large-scale multiplex serology panel was applied in a subset of the UK Biobank (UKB) ^12^, providing a valuable resource to assess pathogen-disease relationships. This serology panel simultaneously measures quantitative antibody responses against antigens of numerous pathogens selected due to their relevance to public health, including human herpesviruses, polyomaviruses, papillomaviruses, and C.trachomatis, H.pylori and T.gondii. Serostatus to these pathogens (i.e. whether or not an individual has been previously infected) can be subsequently derived using seropositivity thresholds, providing a summary of individual infection history. We have since applied this panel to two additional population-based cohorts in the UK, the MRC National Survey of Health and Development (NSHD) ^13^ and Southall and Brent Revisited (SABRE) ^14^, permitting a wide range of pathogens to be investigated in relation to outcomes among several settings in parallel.

In this cross-cohort research, we investigated associations between antibodies to common infections and three established neuroimaging markers of brain structure and pathology with relevance to subclinical dementia – hippocampal volume, whole brain volume (as markers of neurodegenerative disease, particularly Alzheimer’s disease), and white matter hyperintensity volume (as a marker of cerebral small vessel disease). We first examined whether seropositivity and cumulative exposure to 17 pathogens associated with neuroimaging outcomes, and further tested for interactions of these associations with APOE genotype. We then assessed associations of antibody levels against each pathogen (indicative of recent infection or reactivation) with the same neuroimaging outcomes.

## 1. 2. Methods

### 2.1 Study design

An overview of the study design is summarised in Figure 1. Analyses were conducted across three population-based cohorts: 1) Insight 46 ^15^, the neuroscience sub-study of the NSHD – a birth cohort originally consisting of 5362 individuals born in mainland Britain during one week of March 1946 ^13^, 2) SABRE, a tri-ethnic (European, South Asian, and African Caribbean) study consisting of 4972 individuals aged 40-69 at recruitment (1988-1991), randomly selected through GP practices and workplaces in two London boroughs, stratified by ethnicity, sex, and age ^14, 16^, and 3) UKB, a study comprised of >500,000 individuals approximately aged 40-69 at recruitment (2006-2010) ^17^. A summary of study participants can be found in Supplementary Figure 1.

**Figure 1.**
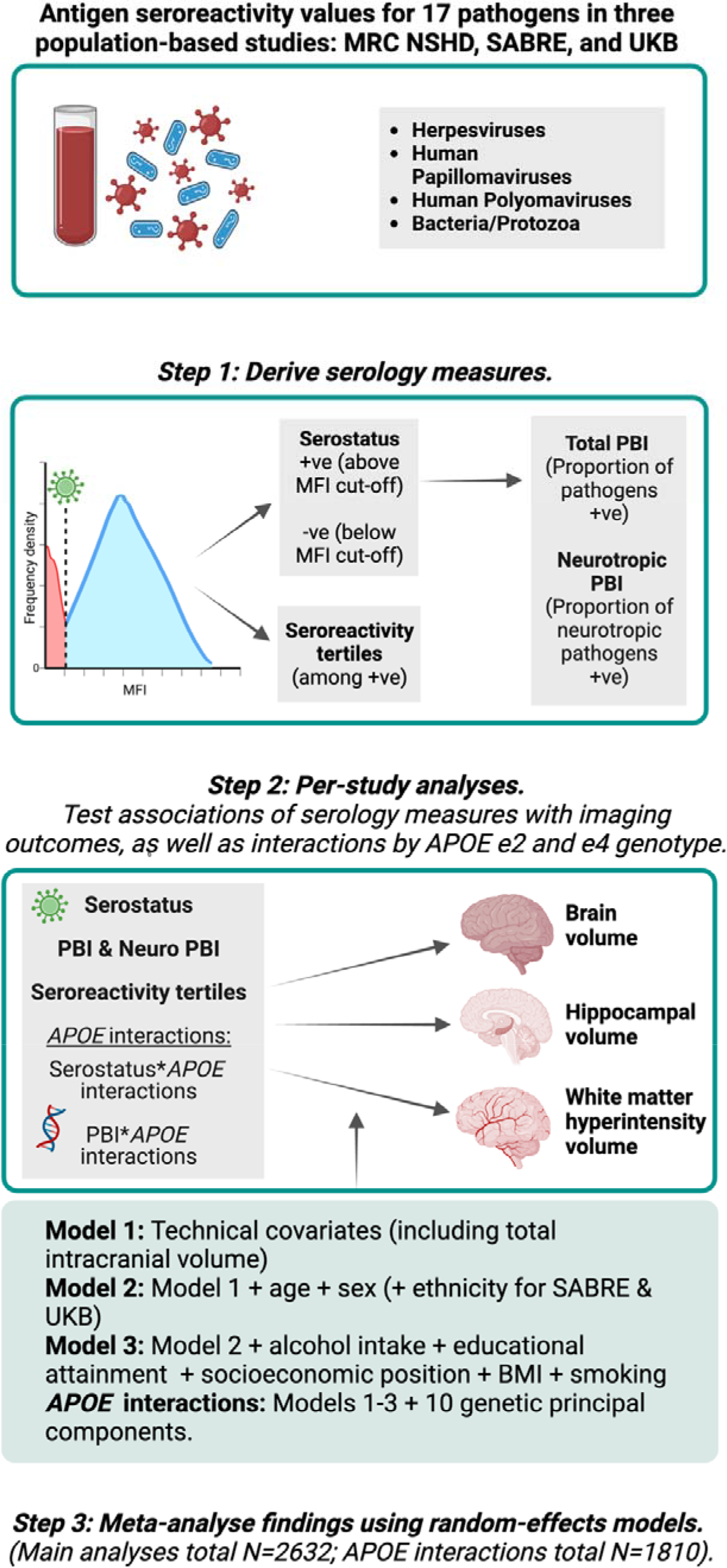
Study workflow. Abbreviations: APOE=Apolipoprotein E genotype; MFI=median fluorescence intensity; MRC NSHD=Medical Research Council National Survey of Health and Development, PBI=pathogen.

### 2.2 Ethical approval

All cohorts received ethical approval and all participants provided written informed consent. The NSHD and its sub-study Insight 46 received approval from the National Research Ethics Service Committee London (14/LO/1173), SABRE from the St Mary’s Hospital Research Ethics Committee (07/H0712/109), and UKB from the NHS North West Research Ethics Committee (11/NW/0382).

### 2.3 Exposure data

#### 2.3.1 Multiplex serology

Serum immunoglobulin G (IgG) antibody levels against a selection of antigens were measured using a validated fluorescence bead-based multiplex serology platform developed at the German Cancer Research Center (DKFZ) in Heidelberg ^12, 18^. Antibody responses (“seroreactivity”) were quantified in median fluorescence intensity units, and between one and six antigens were quantified per pathogen. A full list of pathogens assayed in each cohort are detailed in Supplementary Notes. In UKB, 21 pathogens were assayed among 9429 participants (baseline, 2006-10) and an additional 260 at follow-up only (Instance 1, 2012-13). An adaptation of the same multiplex panel assayed 18 pathogens among 1813 NSHD participants (Age 60-64 follow-up; 2006-10), and 1423 SABRE participants (Visit 2; 2008-11). Thirty-three participants in the NSHD and SABRE were excluded due to technical failures in the assaying (including pipetting errors, high background, and insufficient beadcounts).

#### 2.3.2 Pathogen serostatus and burden scores

We derived binary serostatus variables indicating previous infection for each pathogen, using standardised median fluorescence intensity cut-offs for specific antigens, or antigen combinations where multiple antigens were quantified (see Supplementary Notes). Serostatus variables were derived in the UKB as reported previously ^12^. Only the 17 pathogens with relevant serology data available in all three cohorts were included in this study. To measure cumulative exposure to multiple pathogens, we additionally derived two pathogen burden index (PBI) scores: i) a proportion of positive serostatus values to all 17 pathogens (total PBI; sum of serostatus to 17 pathogens / 17), ii) a proportion of positive serostatus values to 11 neurotropic pathogens, including herpesviruses, T.gondii, and JC virus ^19^ (neurotropic PBI; sum of serostatus to neurotropic pathogens / 11), which may hold more relevance to neurological phenotypes.

#### 2.3.3 Antibody responses

Seroreactivity values formed a variety of non-normal distributions. Therefore, to group antibody responses for seroreactivity analyses, we derived tertiles of antibody titres against each antigen using recommended antigens by the German Cancer Research Center (see Supplementary Notes). Tertiles were derived in the full serology and neuroimaging sample that were seropositive for that antigen.

### 2.4 Neuroimaging outcomes

For NSHD, we used neuroimaging measures collected in the neuroscience sub-study Insight 46, when participants were aged 69-71 years (5-11 years after blood sampling used for serology assaying), using a single Biograph mMR 3 Tesla PET/MRI scanner (Siemens Healthcare) ^15^. For SABRE, we used neuroimaging measures collected at the same time point as blood sampling used for serology assaying (Visit 2). Brain MRI was performed using General Electric Signa HDxt 1.5T, Signa HDx 1.5T, Signa EXCITE 1.5T, and Discovery MR750 3T scanners. Finally, for UKB, neuroimaging measures were collected at the first imaging visit (Instance 2; 1-13 years after blood sampling used for serology assaying). Details of the acquisition protocols for T1 and FLAIR sequences are available in Supplementary Notes.

In all cohorts, brain volume and hippocampal volume were quantified in-house using Geodesic Information Flows ^20^. White matter hyperintensities were derived from T2 FLAIR sequences using an unsupervised automatic algorithm Bayesian Model Selection (BaMoS) (see ^21, 22^ for further details), and were subsequently log_e_ transformed due to skewed distributions.

### 2.5 Genetic data for APOE analyses 2.5.1 Genetic principal components

We used directly genotyped data for genetic quality control and to derive genetic principal components. Details of genotyping can be found for NSHD ^23^, SABRE ^14^, and UKB ^24^ elsewhere. Briefly, blood samples were collected at the age 53 visit for NSHD and baseline (visit 1) for SABRE, and participants were genotyped using the DrugDev array. Two genotyping arrays (BiLEVE Axiom and Affymetrix UK Biobank Axiom) were used to genotype UKB participants. NSHD and SABRE genetic quality control (QC) steps were performed in-house, and we used UKB provided QC criteria and genetic principal components (following the cohort’s central QC) for UKB. For SABRE, QC steps were applied separately by self-reported ethnicity. QC for all cohorts included filtering for common biallelic autosomal variants that did not exhibit significant missingness or deviate from Hardy-Weinberg equilibrium. Samples that did not exhibit significant heterozygosity or missingness, with concordant genetic and self-reported sex, and that were unrelated and closely clustered with reference panel populations using genetic principal components, were retained (see Supplementary Notes for full QC parameters per cohort). Genetic principal components were generated in each study population following pruning and removal of variants in long range linkage disequilibrium regions ^25^. QC steps were performed using plink1.9 and plink2 ^26^, and principal components for NSHD and SABRE were generated using smartpca in EIGENSOFT ^27^.

#### 2.5.2 Apolipoprotein E genotype

APOE genotypes were derived using rs7412 and rs429358 single nucleotide polymorphisms. We subsequently generated binary APOE e4 non-carrier/carrier and APOE e2 non carrier/carrier variables. Carriers were defined as heterozygous or homozygous for the e4 or e2 allele, respectively.

### 2.6 Covariates

Covariates included total intracranial volume and other technical variables related to neuroimaging or serology data collection (NSHD: blood clinic; SABRE: scanner; UKB: scanner co-ordinates and clinic), age at serology and neuroimaging (if not collected at the same time point), sex, ethnicity (SABRE: African-Caribbean, European, South Asian; UKB: Asian or Asian British, Black or Black British, Chinese, Mixed, Other, White), highest educational qualification (none; up to ordinary (‘O’) level or equivalent; advanced (‘A’) level or equivalent, or higher), socioeconomic position (NSHD and SABRE: current or last known occupation categorised into six groups according to the UK Registrar General; UKB: Townsend deprivation index quintiles), BMI (kg/m^2^), smoking status (never, ever), and alcohol intake (low <7 units per week, medium 7-14 units per week, high >14 units per week). A directed acyclic graph detailing the covariate choice can be found in Supplementary Notes.

### 2.7 Statistical analyses

Due to differences in exposure and outcome measurements across studies, data were not pooled and analyses were run in each individual study prior to meta-analysis. In each study, we imputed missing covariate data using multiple imputation by chained equations (10 iterations, 10 datasets; see Table 1 for details on missingness). Analyses were performed on each imputed data set and pooled using Rubin’s rules ^28^. Regression assumptions were checked by examination of the residuals. We meta-analysed findings across studies (or subgroups for APOE interaction analyses, see 2.7.2) using random-effects models with a restricted maximum likelihood estimator. Regression coefficients for analyses with white matter lesion volumes as an outcome were multiplied by 100 to be transformed to sympercents ^29^. We used an exploratory framework for the present analyses, where multiple pathogens were tested in relation to outcomes. Due to these multiple tests, findings from meta-analyses (per outcome for serostatus, seroreactivity, and APOE interaction analyses) were corrected for the false discovery rate (FDR), using the Benjamini-Hochberg procedure ^30^ with an alpha of 0.05. However, given that multiple testing correction increases the likelihood of false negative results, suggestive findings (at unadjusted p<0.05) were additionally reported. Heterogeneity was assessed through Cochran’s Q and I^2^ statistics, and we defined significant heterogeneity where I^2^>50% or Q-p value<0.05. All analyses were performed using R, version 3.6.2.

**Table 1.**
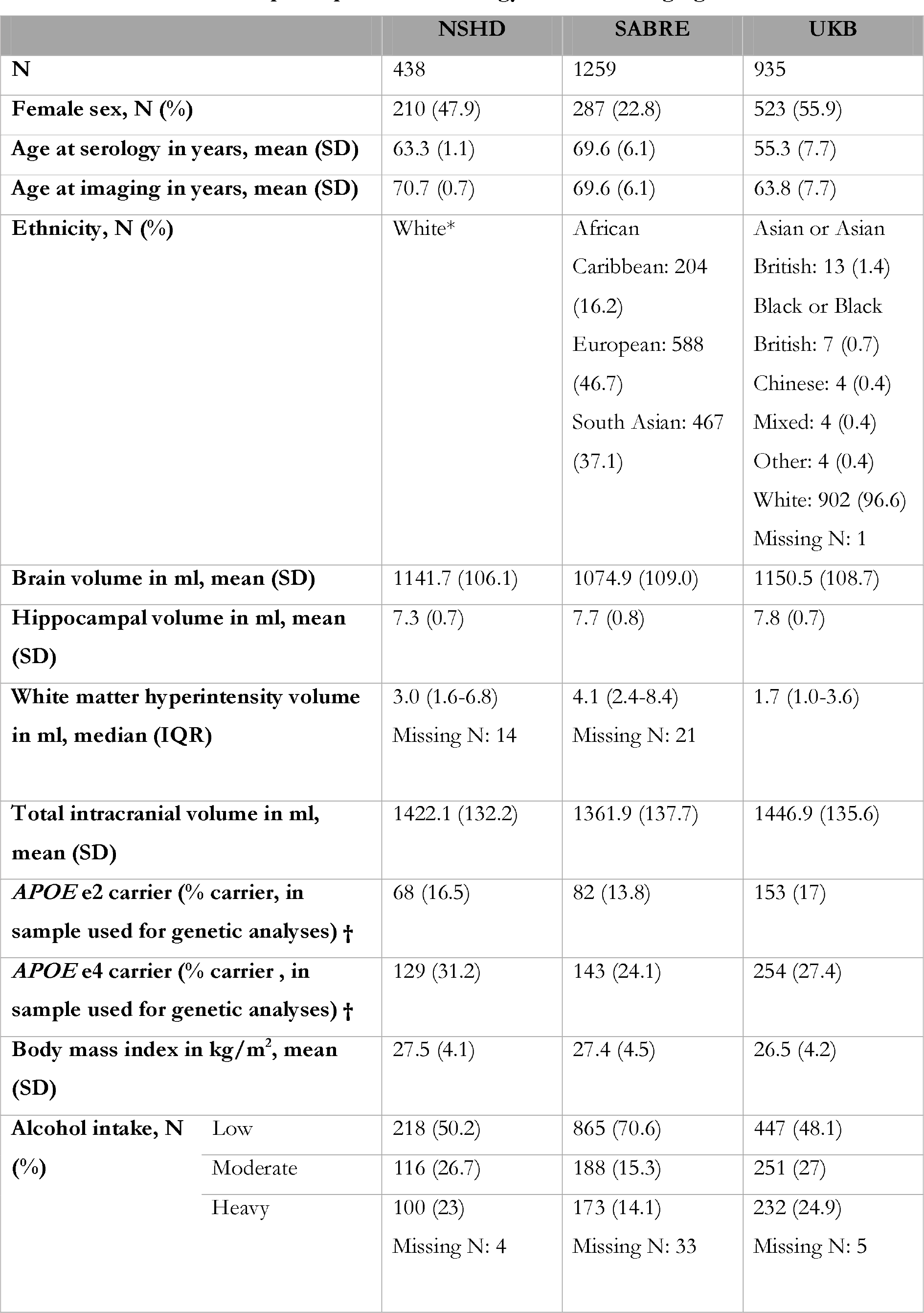
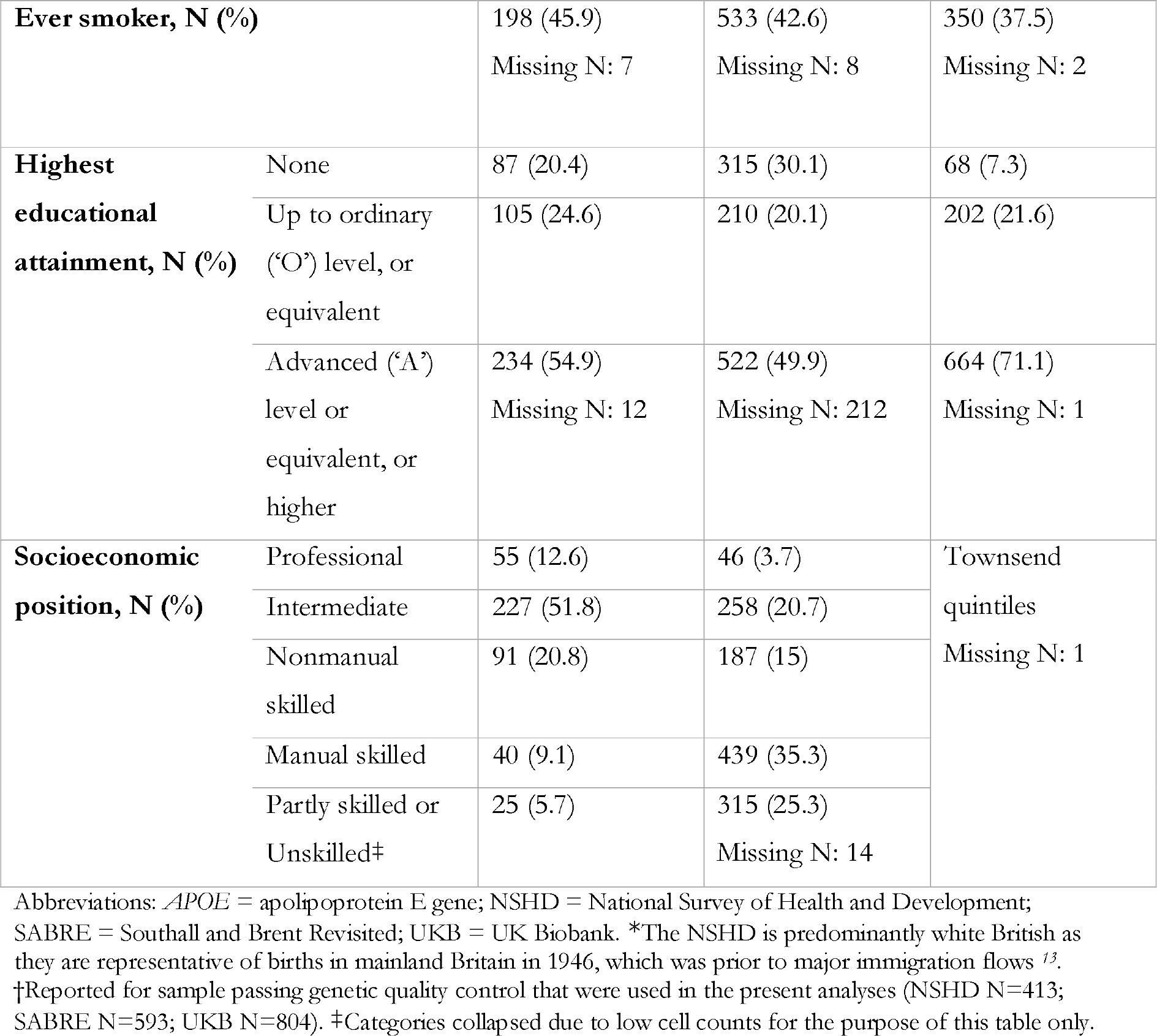
Characteristics of participants with serology and neuroimaging data.

#### 2.7.1 Primary serology analyses

We applied multiple linear regression models to test associations between serology variables (pathogen serostatus and seroreactivity tertiles) and neuroimaging outcomes. Model 1 was minimally adjusted, and included only total intracranial volume and any other technical covariates (NSHD: blood clinic; SABRE: scanner; UKB: scanner co-ordinates and clinic). Model 2 adjusted for model 1 covariates plus basic confounders: age, sex, and (for SABRE and UKB) ethnicity. Model 3 adjusted for model 1 and 2 covariates plus additional possible social, behavioural, and lifestyle confounders: BMI, highest educational qualification, socioeconomic position, smoking status, and alcohol intake. We note that primary exposure for many of these pathogens occurs earlier in life, and thus we included measures reflecting long-term characteristic differences or that may track over time. Statistical models included sampling weights where present. In seroreactivity analyses, tertiles were modelled as an ordinal variable, and pathogens with a seroprevalence over 5% in all studies were investigated (the two Human Papillomaviruses (HPV), and Kaposi’s sarcoma-associated herpesvirus (KSHV) were therefore not studied). For pathogens where multiple antigens were quantified, we randomly selected one from the subset of recommended antigens (see Supplementary Notes) using the sample function in base R, with the other antigens investigated in sensitivity analyses.

#### 2.7.2 APOE interaction analyses

To examine whether serostatus or pathogen burden relationships with outcomes differed by APOE genotype, we performed the same nested models listed above including a) APOE e4 carrier status, and b) APOE e2 carrier status as an interaction term. APOE e4 and APOE e2 analyses were conducted separately. All models were additionally adjusted for 10 genetic principal components, a recommended practice to minimise potential confounding by population structure, and for SABRE, analyses were further stratified by European and South Asian groups in the sample (sample size was not sufficient for inclusion of the African Caribbean group, N=22). Coefficients for interaction terms from each analysis were then meta-analysed using random-effects models as described in 2.7. Due to sample size (and hence power) limitations, we did not test APOE interactions for seroreactivity tertile associations with outcomes, which include subsets of participants seropositive to the antigen of interest. Where suggestive evidence (unadjusted p<0.05) of an interaction was observed, analyses were rerun stratified by APOE genotype.

### 2.8 Secondary and sensitivity analyses

We conducted several additional analyses, as follows: 1) we repeated serostatus analyses using alternate serostatus definitions for three pathogens (HPV16, C.trachomatis, H.pylori) that were available in the UKB; 2) for pathogens with multiple antigens assayed, we repeated seroreactivity analyses using alternate antigens, as detailed in 2.3.3; 3) we restricted samples to participants who were ≥60 years at the time of blood sampling used for serology assays, and performed the same analytical models as detailed above (NSHD N=438 – all aged 60-64; SABRE N=1240; UKB N= 331); 4) we repeated all primary analyses excluding participants with a diagnosis of dementia or stroke (to include participants with only subclinical structural and cerebrovascular pathology only), or with known conditions affecting immune function (of which only diabetes was available in all cohorts, see Supplementary Notes), defined at or before the time of blood sampling used for serology assays (NSHD N=408; SABRE N=831; UKB N=901).

## 1. 3. Results

### 3.1 Cohort characteristics and seroprevalence

Participant characteristics can be found in Table 1. Characteristics and seroprevalences for the full serology cohort and the serology and neuroimaging subset can be found in Supplementary Table 1. In the present study, we included participants with available serology measures and with data on at least one neuroimaging outcome (Total N: 2632; NSHD N=438, SABRE N=1259, UKB N=935). For APOE genotype interactions, available genetic data following QC were additionally required (Total N: 1810; NSHD N=413; SABRE = 314 (European), 279 (South Asian); UKB=804).

### 3.2 Serostatus and pathogen burden

Meta-analysed findings for serostatus and PBI analyses are displayed in Figure 2 and Table 2 (see Supplementary Figures 2-4 for per cohort findings, and Supplementary Tables 2-4 for full numerical results). To give magnitudes of associations context, a 1-year increase in age has been associated with a 16.5% increase in white matter lesions, 11.5ml smaller whole brain volume, and 0.08ml smaller hippocampal volume in the NSHD between 69-71 years of age ^31^.

**Figure 2.**
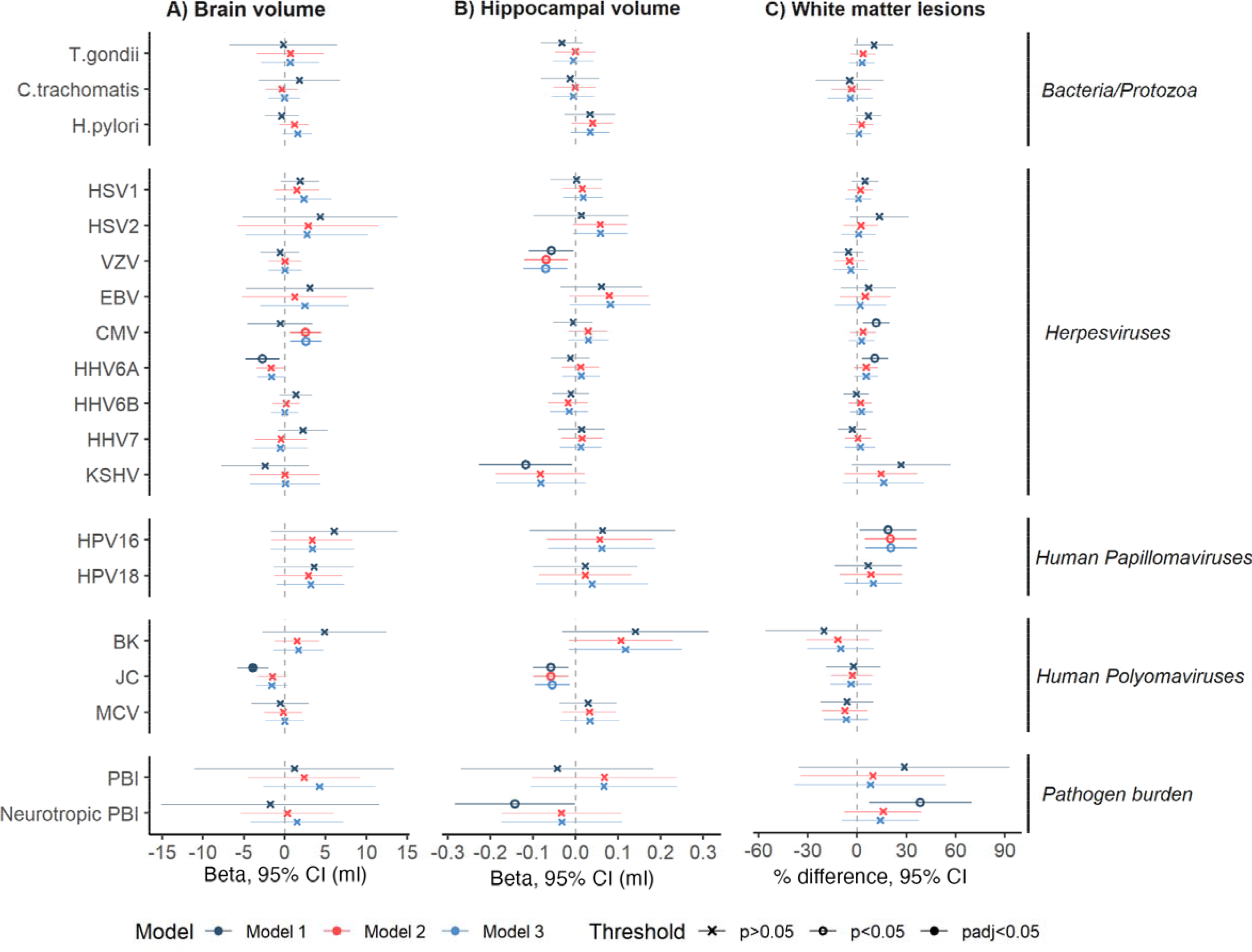
(left). Forest plot indicating meta-analysed associations of pathogen serostatus and burden scores neuroimaging outcomes. Positive estimates indicate large brain and hippocampal volume more white matter lesions. For pathogen burden analyses, estim represent the change in outcom 0.1 unit increase in pathogen bu (proportion of pathogens or neurotropic pathogens seropos to). Model 1 included adjustme for total intracranial volume an other technical covariates; mod additionally for age, sex, and ethnicity; and model 3 addition for BMI, smoking status, educa attainment, socioeconomic pos and alcohol intake. Statistical m are indicated by colour, and significance threshold by shape Pathogen families are further indicated on the right-hand side the figure. Abbreviations: BK=BK virus; CMV=Cytomegalovirus; EBV=Eps Barr virus; HHV=Human herpesvir HPV=Human Papillomavirus; HSV=Herpes simplex virus; JC=Jo Cunningham virus; KSHV=Kaposi’ sarcoma-associated herpesviruses; MCV=Merkel Cell virus; PBI=Path burden index; VZV=Varicella zoste virus.

**Table 2.** Meta-analysed associations of serostatus and pathogen burden scores with neuroimaging outcomes. Estimates, 95% confidence intervals, and p values are presented. For pathogen burden analyses, estimates represent the difference in outcome per 0.1 unit inc rease in pathogen burden (proportion of pathogens or neurotropic pathogens seropositive to). Associations significant at p<0.05 are in bold, but none pr esented survived multiple testing correction. Significant heterogeneity (either I statistic >50 and/or Cochran’s Q p<0.05) are indicated by †. Due to space, only results from model 2 (adjusting for total intracranial volume and other technical covariates, age, sex, and ethnicity) and model 3 (additionally adjusting for BM I, smoking status, educational attainment, socioeconomic position, and alcohol intake) are shown.

|  | Brain volume |  | Hippocampal volume |  | White matter lesions |  |
| --- | --- | --- | --- | --- | --- | --- |
|  | Model 2 | Model 3 | Model 2 | Model 3 | Model 2 | Model 3 |
| | $\beta$ (ml) [95% CI] | $\beta$ (ml) [95% CI] | $\beta$ (ml) [95% CI] | $\beta$ (ml) [95% CI] | % change [95%CI] | % change [95%CI] |
| HSV1 | 1.48 [-1.26,4.23]† | 2.35 [-1.04,5.73]† | 0.02 [-0.03,0.06] | 0.02 [-0.03,0.06] | 2.08 [-5.47,9.62] | 0.8 [-6.88,8.47] |
| HSV2 | 2.87 [-5.81,11.54]† | 2.73 [-4.73,10.19]† | 0.06 [-0.01,0.12] | 0.06 [0,0.12] | 2.36 [-8.02,12.74] | 0.94 [-9.47,11.35] |
| VZV | 0.03 [-2,2.07] | 0.04 [-1.97,2.06] | **-0.07 [-0.12,-0.02]** | **-0.07 [-0.12,-0.02]** | -4.47 [-13.63,4.7] | -3.82 [-14.6,6.96] |
| EBV | 1.21 [-5.19,7.62]† | 2.45 [-2.98,7.88] | 0.08 [-0.01,0.17] | 0.08 [-0.01,0.18] | 5.03 [-10.45,20.51] | 2 [-13.66,17.66] |
| CMV | **2.57 [0.66,4.48]** | **2.62 [0.71,4.53]** | 0.03 [-0.02,0.08] | 0.03 [-0.02,0.08] | 3.61 [-4.15,11.36] | 2.84 [-4.98,10.67] |
| HHV6A | -1.68 [-3.45,0.09] | -1.61 [-3.37,0.15] | 0.01 [-0.03,0.06] | 0.01 [-0.03,0.06] | 5.5 [-1.77,12.77] | 5.48 [-1.81,12.76] |
| HHV6B | 0.17 [-1.52,1.85] | 0 [-1.68,1.67] | -0.02 [-0.06,0.03] | -0.01 [-0.06,0.03] | 2.06 [-4.86,8.97] | 2.79 [-4.17,9.75] |
| HHV7 | -0.46 [-3.62,2.71] | -0.56 [-3.99,2.86] | 0.02 [-0.03,0.06] | 0.01 [-0.04,0.06] | 0.52 [-7.39,8.43] | 2.02 [-7.25,11.29] |
| KSHV | 0.02 [-4.26,4.31] | 0.07 [-4.21,4.35] | -0.08 [-0.19,0.02] | -0.08 [-0.19,0.02] | 14.61 [-7.55,36.77] | 16.14 [-8.48,40.77] |
| HPV16 | 3.35 [-1.6,8.29] | 3.37 [-1.76,8.51] | 0.06 [-0.07,0.18] | 0.06 [-0.06,0.19] | **20.48 [4.77,36.18]** | **20.78 [5.03,36.53]** |
| HPV18 | 2.9 [-1.27,7.07] | 3.18 [-0.97,7.32] | 0.02 [-0.09,0.13] | 0.04 [-0.09,0.17] | 8.48 [-10.49,27.46] | 9.93 [-7.48,27.33] |
| BK | 1.5 [-1.22,4.23] | 1.69 [-1.37,4.74] | 0.11 [-0.01,0.23]† | 0.12 [-0.01,0.25]† | -11.77 [-30.99,7.45] | -10.03 [-30.32,10.26] † |
| JC | -1.51 [-3.31,0.29] | -1.59 [-3.55,0.36] | **-0.06 [-0.1,-0.02]** | **-0.05 [-0.09,-0.01]** | -3 [-15.5,9.5]† | -3.64 [-16.18,8.9]† |
| MCV | -0.16 [-2.47,2.14] | 0 [-2.38,2.37] | 0.03 [-0.03,0.1]† | 0.03 [-0.04,0.1]† | -7.42 [-21.24,6.4]† | -6.58 [-20.09,6.93]† |
| Ct | -0.35 [-2.32,1.61] | -0.03 [-1.98,1.93] | 0 [-0.05,0.05] | 0 [-0.05,0.05] | -3.39 [-15.33,8.54] | -4.07 [-17.81,9.67] |
| Tg | 0.69 [-3.42,4.8]† | 0.67 [-2.88,4.22]† | 0 [-0.05,0.05] | 0 [-0.05,0.04] | 3.67 [-4.19,11.52] | 3.04 [-4.84,10.92] |
| Hp | 1.18 [-0.61,2.96] | 1.59 [-0.2,3.38] | 0.04 [-0.01,0.09] | 0.03 [-0.01,0.08] | 2.76 [-4.58,10.09] | 1.1 [-6.33,8.52] |
| **PBI** | 0.24 [-0.45,0.92] | 0.42 [-0.26,1.11] | 0.01 [-0.01,0.02] | 0.01 [-0.01,0.02] | 0.96 [-3.42,5.34]† | 0.82 [-3.81,5.44]† |
| **Neuro PBI** | 0.03 [-0.54,0.6] | 0.15 [-0.42,0.72] | 0 [-0.02,0.01] | 0 [-0.02,0.01] | 1.58 [-0.75,3.91] | 1.41 [-0.93,3.76] |

Abbreviations: BK=BK virus; CMV=Cytomegalovirus; Ct=*C.trachomatis*; EBV=Epstein-Barr virus; HHV=Human herpesvirus; Hp=*H.pylori*; HPV=Human Papillomavirus; HSV=Herpes simplex virus; JC=John Cunningham virus; KSHV=Kaposi's sarcoma-associated herpesviruses; MCV=Merkel Cell virus; PBI=Pathogen

Seropositivity to JC virus was associated with smaller brain volume in model 1 (brain volume, ß=-3.89ml [-5.81,-1.97], p=7.3×10^-5^, p_FDR_<0.05). We observed a notable attenuation following model 2 adjustments (age, sex, and ethnicity where relevant) and negligible further changes in the fully adjusted model (ß=-1.5ml [-3.55,0.36], p=0.11). We found no clear evidence of associations between other pathogens or PBI and outcomes after multiple testing correction, although four further suggestive relationships were observed in fully adjusted models (seropositivity to cytomegalovirus (CMV) with larger brain volume, varicella zoster virus (VZV) and JC virus with smaller hippocampal volume, and HPV16 and more white matter lesions). There were also some inconclusive results with potentially noteworthy point estimates but wide 95% confidence intervals that crossed the null, such as KSHV in association with more white matter lesions. The pattern of results did not differ meaningfully in our sensitivity analyses (see Supplementary Figures 5-7).

#### 3.2.1 APOE interactions

Meta-analysed findings for APOE interactions are displayed in Supplementary Table 5. We found no evidence of interactions by APOE e4 or APOE e2 carrier status following multiple testing correction. In model 3, we observed suggestive evidence of an interaction between APOE e2 carrier status and four serology measures (seropositivity to T.gondii and MCV, and both the PBI and neurotropic PBI) with smaller hippocampal volume. We subsequently tested associations of these serology measures with hippocampal volume stratified by APOE e2 carrier status. Results from all models can be found in Supplementary Table 6. In the final model, we report associations between PBI and smaller hippocampal volume among APOE e2 carriers only (per 10% increase in PBI ß=-0.07ml,[-0.13,-0.01], p=0.01).

### 3.3 Seroreactivity tertiles

Meta-analysed findings for seroreactivity analyses are displayed in Figure 3 and Table 3 (see Supplementary Figures 8-10 for study-level findings, and Supplementary Tables 7-9 for full numerical results).

**Figure 3.**
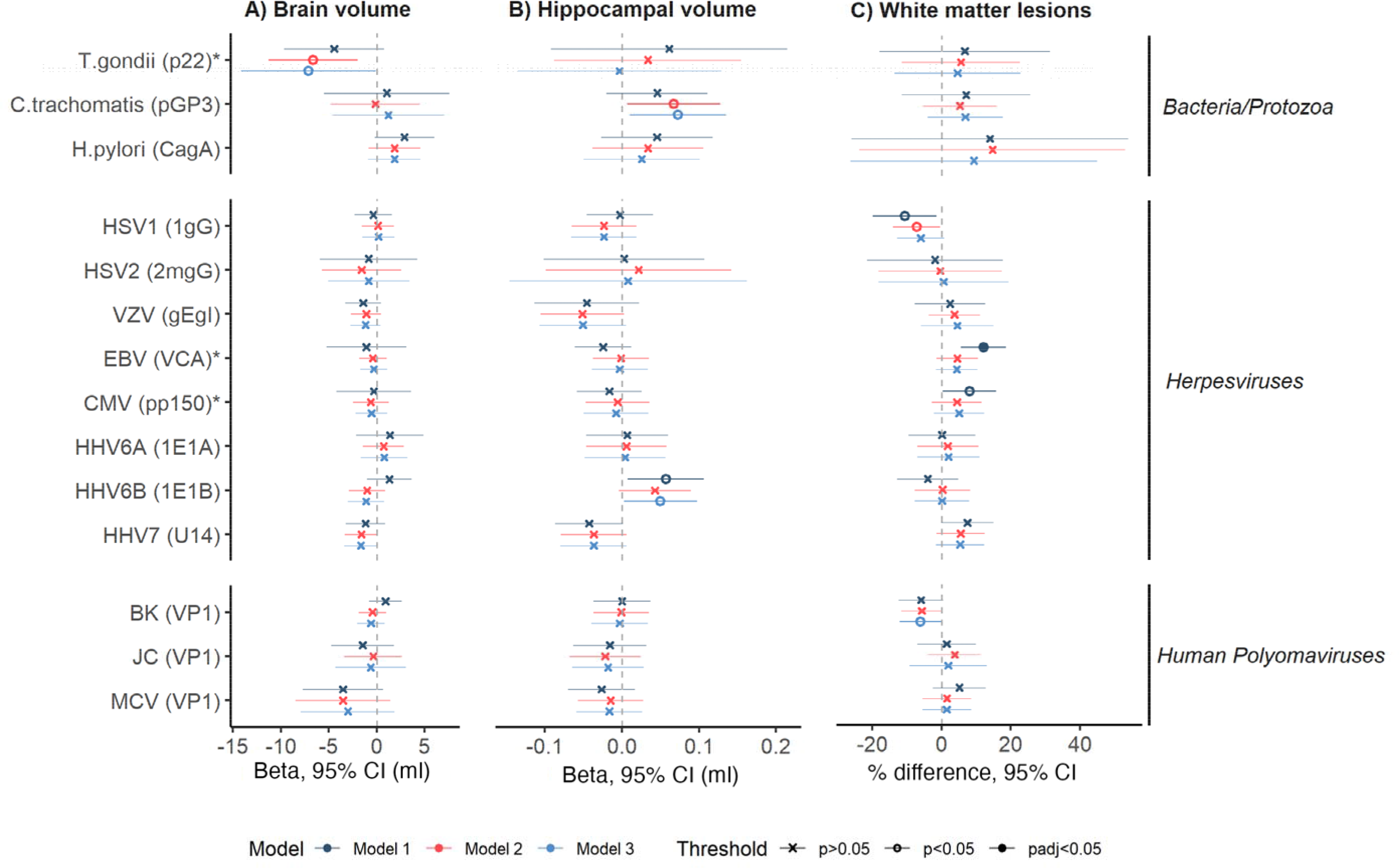
Forest plot indicating meta-analysed results between seroreactivity tertiles and neuroimaging outcomes. Positive estimates indicate larger brain and hippocampal volumes, but more white matter lesions. Model 1 included adjustments for total intracranial volume and other technical covariates; model 2 additionally for age, sex, and ethnicity; and model 3 additionally for BMI, smoking status, educational attainment, socioeconomic position, and alcohol intake. Statistical models are indicated by colour, and significance threshold by shape. Pathogens and specific antigens (in brackets) are indicated on the y-axis, with pathogen families further indicated on the right-hand side of the figure. Antigens selected at random from a larger p ool of recommended antigens for the pathogen are indicated by an asterisk. Abbreviations: BK=BK virus; CMV=Cytomegalovirus; EBV=Epstein-Barr virus; HHV=Human herpesvirus; HPV=Human Papillomavirus; HSV=Herpes simplex virus; JC=John Cunningham virus; MCV=Merkel Cell virus; VZV=Varicella zoster virus.

**Table 3.** Meta-analysed associations of seroreactivity tertiles with neuroimaging outcomes. Estimates, 95% confidence intervals, and p values are presented. Associations significant at p<0.05 are in bold, but none presented survived multiple testing correction. Significant heterogeneity (either I statistic >50 and/or Cochran’s Q p<0.05) are indicated by †. Due to space, only results from model 2 (adjusting for total intracranial volume, other technical covariates, age, sex, and ethnicity) and model 3 (additionally adjusting for BMI, smoking status, educational attainment, socioeconomic position, and alcohol intake) are shown.

|  | Brain volume |  | Hippocampal volume |  | White matter lesions |  |
| --- | --- | --- | --- | --- | --- | --- |
|  | Model 2 | Model 3 | Model 2 | Model 3 | Model 2 | Model 3 |
| | $\beta$ (ml)[95% CI] | $\beta$ (ml)[95% CI] | $\beta$ (ml)[95% CI] | $\beta$ (ml)<br>[95% CI] | % change [95%CI] | % change [95%CI] |
| **HSV1** | 0.12 [-1.58,1.81] | 0.18 [-1.52,1.87] | -0.02[-0.07,0.02] | -0.02[-0.07,0.02] | **-7.19[-13.99,-0.39]** | -6.01[-12.83,0.82] |
| **HSV2** | -1.58 [-5.7,2.54] | -0.85 [-5.1,3.41] | 0.02 [-0.1,0.14] | 0.01[-0.15,0.16] | -0.41[-18.24,17.43] | 0.56[-18.21,19.32] |
| **VZV** | -1.1 [-2.67,0.47] | -1.18 [-2.74,0.39] | -0.05[-0.11,0] | -0.05[-0.11,0.01] | 3.7[-3.67,11.07] | 4.53[-5.93,14.99]† |
| **EBV** | -0.41 [-1.84,1.03] | -0.31 [-1.73,1.12] | 0 [-0.04,0.03] | 0 [-0.04,0.03] | 4.58[-1.36,10.52] | 4.44[-1.53,10.4] |
| **CMV** | -0.62 [-2.5,1.25] | -0.55 [-2.23,1.12] | -0.01[-0.05,0.04] | -0.01[-0.05,0.03] | 4.46[-2.75,11.66] | 5.05[-2.21,12.3] |
| **HHV6A** | 0.71 [-1.42,2.83] | 0.77 [-1.68,3.21] | 0.01 [-0.05,0.06] | 0 [-0.05,0.06] | 1.82[-7.02,10.66] | 1.99[-6.99,10.98] |
| **HHV6B** | -1.02 [-2.93,0.88] | -1.12 [-3.01,0.78] | 0.04[0,0.09] | **0.05 [0,0.1]** | 0.33[-7.7,8.36] | 0.18[-7.71,8.07] |
| **HHV7** | -1.62 [-3.35,0.12] | -1.65 [-3.38,0.07] | -0.04[-0.08,0.01] | -0.04[-0.08,0.01] | 5.52[-1.46,12.49] | 5.37[-1.65,12.39] |
| **BK** | -0.45 [-1.9,1] | -0.59 [-2.04,0.85] | 0 [-0.04,0.03] | 0 [-0.04,0.03] | -5.67[-11.61,0.28] | **-6.1[-12.08,-0.11]** |
| **JC** | -0.37 [-3.37,2.63]† | -0.63 [-4.33,3.07]† | -0.02[-0.07,0.02] | -0.02[-0.06,0.03] | 3.78[-3.76,11.31] | 1.95[-9.23,13.13] |
| **MCV** | -3.52 [-8.45,1.41]† | -3.02 [-7.93,1.88]† | -0.01[-0.06,0.03] | -0.02[-0.06,0.03] | 1.56[-5.41,8.54] | 1.52[-5.52,8.55] |
| **Ct** | -0.14 [-4.81,4.52] | 1.2 [-4.64,7.04]† | **0.07 [0.01,0.13]** | **0.07[0.01,0.14]** | 5.33[-5.33,15.99] | 6.87[-4.04,17.78] |
| **Tg** | **-6.63 [-11.26,-2]** | **-7.11 [-14.13,-0.08]** | 0.03[-0.09,0.15] | 0[-0.14,0.13] | 5.58[-11.46,22.61] | 4.63[-13.49,22.75] |
| **Hp** | 1.86 [-0.84,4.55] | 1.84 [-0.9,4.59] | 0.03[-0.04,0.11] | 0.03[-0.05,0.1] | 14.71[-23.7,53.12]† | 9.36[-26.28,44.99]† |

Abbreviations: BK=BK virus; CMV=Cytomegalovirus; Ct=*C. trachomatis*; EBV=Epstein-Barr virus; HHV=Human herpesvirus; Hp=*H. pylori*; HSV=Herpes simplex virus; JC=John Cunningham virus; MCV=Merkel Cell virus; Tg=*T. gondii*; VZV=Varicella zoster virus. Kaposi's sarcoma-associated herpesviruses and Human Papillomavirus 16 and 18 were not tested as seroprevalence were $< 5\%$ in at least one cohort.

Seroreactivity to Epstein-Barr virus (EBV; VCA antigen) was associated with more white matter lesions in model 1 (12.1% difference [5.7, 18.6], p=2.3×10^-4^, p _FDR_<0.05) but largely attenuated in subsequent models (model 3: 4.5% difference [-1.5,10.1], p=0.15). We additionally found less evidence of an association between seroreactivity to the alternate EBV antigen (EBNA) and white matter lesions in sensitivity analyses (model 1: 4.6% difference [-9.6, 18.8], p=0.53). We found no further clear evidence of associations between the remaining antigens and outcomes following multiple testing correction, although other suggestive associations were observed in fully adjusted models (T.gondii with smaller brain volume (which was not robust when testing the alternate antigen); human herpesvirus (HHV)6B and C.trachomatis with larger hippocampal volume, and BK virus with less white matter lesions). With the exception of the T.gondii results reported above, the pattern of results did not differ appreciably when running sensitivity analyses (see Supplementary Figures 11-13).

## 1.4. Discussion

In this cross-cohort study using three population-based cohorts, we examined associations of multiple common pathogens with neuroimaging markers related to subclinical dementia – brain volume, hippocampal volume, and white matter lesions – and tested whether these associations were modified by APOE e4 and e2 carrier status. We found little or no evidence of strong relationships in most instances. Nevertheless, several suggestive associations require examination in other cohorts with serology and neuroimaging data, given the possibility of non-negligible magnitudes of association – e.g. the difference in hippocampal volume between VZV-seropositive and seronegative individuals (−0.07ml) equates to approximately two thirds of the life course-averaged effect of APOE ε4 carriage on hippocampal volume (−0.11ml) ^32^.

Among infectious agents that may have a role in the aetiology of dementia, links between HSV and Alzheimer’s disease (AD) are perhaps the most studied to date ^33^. Many of the other pathogens with serological markers on our panel have also been associated with AD or other causes of dementia, although findings have been contested ^34^. Nevertheless, our null findings for HSV – alongside those for other pathogens – in relation to neuroimaging outcomes are in line with several other studies. For example, serostatuses for C.pneumoniae, H.pylori and CMV were not associated prospectively with dementia risk, whole brain volume, or white matter lesions in the Framingham cohort study ^35^. In a study using data from the Baltimore Longitudinal Study of Ageing, no associations were found between symptomatic herpesvirus infections and changes in brain volume or AD signature regions; although some associations were reported for other markers of dementia risk, including attentional decline and astrogliosis ^36^. Null findings between HSV1 serostatus and whole brain atrophy were additionally reported in a cohort with parental history of early-onset autosomal dominant AD ^37^. In contrast, relationships between higher seroreactivity to HSV (either 1 or 2) and smaller hippocampal volume were reported in an analysis involving two small cohorts (N=349) ^38^. Additionally, using the same multiplex serology data as the present study, an association of HSV1 serostatus with incident dementia diagnosis was observed in the UKB. No relationships were clearly identifiable for the other pathogens; however, with only 84 incident dementia cases available in the sample, that analysis may have lacked power to detect moderate but clinically meaningful associations ^39^.

There could be several explanations where associations of pathogens with neuroimaging measures were not observed. Since severe infections, including those leading to hospitalisation, appear to be risk factors for AD ^40^, virulent infections (rather than any exposure or seroreactivity against pathogen antigens measured by serology) could be more relevant to dementia risk. Alternatively, pathogen exposure might only have a role later in the disease trajectory, rather than in early pathogenesis. Participants included in the present analyses are still relatively young (mean age 63.8-70.7 among the cohorts studied) and could be many years prior to developing overt signs of neurodegeneration and cerebrovascular pathology. A further possibility is that interactions between pathogens, or a combined burden of many, would be necessary to drive the development of neuropathology. In accordance with this hypothesis, pathogen burden indices derived from counts of serostatus values have been associated with dementia risk in other settings ^39, 41^, but we found no convincing evidence of associations of pathogen burden scores with neuroimaging outcomes. Finally, relationships may additionally be contingent on interactions with other environmental or genetic factors. For instance, one study observed increased AD risk with higher HSV1 seroreactivity in APOE e4 carriers only ^9^, and other data suggested that CMV and H.pylori serostatuses associated differently with whole brain volume according to e4 carriage ^35^. We did not find evidence of APOE e4 modification of serostatus associations with neuroimaging measures; however, identifying such interactions robustly in exploratory analyses with many statistical tests like ours may require much larger sample sizes. We found some suggestive evidence for associations of exposures with smaller hippocampal volume in APOE e2 carriers only, but are not aware of any reason why these would be specific to APOE e2. It is possible this is a chance finding, and would want to see whether the results are robust to replication, although we note that a previous study reported an association of increasing pathogen burden with worse cognitive function in non-APOE e4 carriers only ^10^.

Among other suggestive findings that we observed, associations of seropositivity to VZV and JC virus with smaller hippocampal volumes, and of HPV16 seropositivity with more white matter lesions, have plausibility. VZV infection and its clinical manifestation as shingles have been investigated as risk factors for dementia in many settings, although findings have been mixed ^8, 42^. JC virus is able to cause a rare, aggressive neurodegenerative disease under immunosuppressed conditions ^43^, and some evidence has suggested associations of HPV infection with cardiovascular outcomes ^44^. To our knowledge, no other studies have investigated associations of these infections with subclinical neuroimaging outcomes. No prospective associations of serostatus for these pathogens with all-cause dementia risk were detected by a study using UKB data, although, as mentioned, results in this study suffered from a lack of statistical precision ^39^; in particular, their finding for VZV clearly requires follow-up in larger data (OR=3.38 for seropositive individuals; 95% CI: 0.83, 13.78). In contrast, some of our other suggestive findings were unanticipated based on our hypothesis that pathogen exposures would be associated with worse neuroimaging metrics. For example, the associations of CMV seropositivity with larger whole brain volumes, and three of the seroreactivity measures with larger hippocampal volume (for HHV6B and C.trachomatis) or fewer white matter lesions (for BK virus). Possible reasons for these associations are unclear, and we are not aware of other studies reporting relationships of these infections with dementia or related outcomes in this direction. As with any of the suggestive findings reported, these may have arisen due to chance or bias, e.g. from residual confounding, and we stress the need to see whether these results can be replicated in other data.

### Strengths and limitations

This study is the largest and broadest – in terms of number of pathogens examined – to investigate associations of common infections with neuroimaging markers of dementia to date. We used harmonised exposure and neuroimaging data from three large and well-characterised population-based cohorts, with an array of data on possible confounders. Furthermore, multiplex serology data directly ascertains pathogen exposure and circumvents reliance on inaccurate diagnoses from medical records or self-report of symptomatic infections from surveys. The use of subclinical markers of dementia risk are additionally less prone (although not immune) to biases, such as survival bias, in comparison to studies with clinical endpoints as outcomes. We also note several limitations. First, these findings are observational and hence associations might still arise from reverse causality, residual confounding, or other sources of bias. While most of the infections we studied are typically acquired earlier in life and thus are likely to precede the onset of neuropathology, the long prodromes of AD and other causes of dementia may mean neurodegenerative processes could influence immune function and subsequent serological values over long periods. Second, participants remaining in NSHD are only broadly representative of the study population at recruitment and Insight 46 study members are on average of better self-reported health, cognitive function, and socioeconomic position than the full NSHD cohort ^45, 46^. In addition, participants in the UKB (and particularly the imaging subset) are healthier and more socially advantaged than the general UK population at the same age range ^17, 47^. We incorporated sampling weights for NSHD, but no weighting was applied in the other cohorts. Third, serology values were measured once, and some individuals may have been misclassified – particularly in NSHD and UKB if individuals were infected in the period between sample collection for serology assaying and the neuroimaging visits. Fourth, as seroreactivity analyses were conducted in the seropositive subset only, results may be biased through selection on serostatus. Fifth, we were unable to identify the age of infection onset or severity of infection with these data, and therefore investigations that addressed the timing and severity of infections were not possible, although the use of seroreactivity values as exposures may indicate more recent infection or reactivation. Sixth, as primary infections for many of the studied pathogens typically occur earlier in life, some of the behavioural and lifestyle variables included in model 3 may occur following infection and be inappropriate as confounders. Nevertheless, model 3 adjustments did not result in large changes to most results. Finally, we restricted the neuroimaging outcomes to those available in all three cohorts (measures of brain structure and white matter lesions), and thus did not address potential relationships of these exposures with more specific pathologies of dementia subtypes, such as amyloid.

### Future directions

There are several ways to build upon the evidence presented in this study. The generation of equivalent serology data (or other molecular measures of infection), in studies with neuroimaging data and/or clinical follow-up, will help to estimate associations with more precision to establish or refute some of the suggestive findings observed in this study. Given many of these infections are preventable or treatable, this could help inform public health strategies, such as the prioritisation and coverage of vaccination programmes. Furthermore, expanding this work to include longitudinal outcomes, such as rates of atrophy and change in white matter lesion burden could provide more granular insights into relationships with neurodegeneration and cerebrovascular pathology. Finally, incorporating other measures of neuropathology (e.g. cerebral amyloidosis), and fluid-based biomarkers of neurodegeneration will help to triangulate possible effects of pathogens on multiple pathways affecting dementia risk.

## Data Availability

Per-study results are available on request.

## Acknowledgements

We are grateful to study participants of the NSHD (and Insight 46), SABRE, and UKB. We thank Lee Hamill Howes, Andrew Wong, Kenan Direk, and Felicia Huang, as well as other members of the Study Support Team at the MRC Unit for Lifelong Health and Ageing at UCL for their invaluable help towards the generation and curation of NSHD and SABRE data used in this project. We are also grateful to the radiographers and nuclear medicine physicians at the UCL Institute of Nuclear Medicine and to the staff at the Leonard Wolfson Experimental Neurology Centre at UCL. We are particularly indebted to the support of the late Dr Chris Clark of Avid Radiopharmaceuticals who championed the Insight 46 study from its outset. This research has been conducted using the UK Biobank Resource under Application Number 71702. JMS acknowledges the support of the National Institute for Health Research University College London Hospitals Biomedical Research Centre, Wolfson Foundation, Alzheimer’s Research UK, Brain Research UK, Weston Brain Institute, Medical Research Council, British Heart Foundation, UK Dementia Research Institute and Alzheimer’s Association. We further acknowledge the use of BioRender.com for the creation of Figure 1.

## Data availability

Per-study results are available on request. Analytical code will be made publicly available.

## Abbreviations

AD: Alzheimer’s disease
APOE: Apolipoprotein E
BaMoS: Bayesian Model Selection
BMI: Body mass index
CMV: Cytomegalovrius
EBV: Epstein-Barr virus
HHV: Human herpesvirus
HPV: Human papillomavirus
HSV: Herpes simplex virus
IgG: Immunoglobulin G
JC: John-Cunningham virus
KSHV: Kaposi’s sarcoma-associated herpesviruses
MCV: Merkel Cell virus
NSHD: National Survey of Health and Development
PBI: Pathogen burden index
QC: Quality control
SABRE: Southall and Brent Revisited
UKB: UK Biobank
VZV: Varicella zoster virus

