## Supplementary Table for "Common infections and neuroimaging markers of dementia in three UK cohort studies"

Supplementary Tables

**Supplementary Table 1. Characteristics of participants and seroprevalence for the 17 pathogens common to all cohorts.**

| | | **NSHD** | | **SABRE** | | **UKB** | |
|---|---|---|---|---|---|---|---|
| | | *Serology* | *Serology & Imaging* | *Serology* | *Serology & Imaging* | *Serology* | *Serology & Imaging* |
| **N** | | 1793 | 438 | 1410 | 1259 | 9689 | 935 |
| **Sex (% female)** | | 50.5 | 47.9 | 23.7 | 22.8 | 55.9 | 55.9 |
| **Age at serology in years, mean (SD)** | | 63.2 (1.1) | 63.3 (1.1) | 69.6 (6.1) | 69.6 (6.1) | 56.7 (8.2) | 55.3 (7.7) |
| **Herpesviruses** | Herpes simplex virus-1† (% seropositive) | 67.3 | 64.4 | 81.6 | 81.1 | 69.8 | 61.5 |
| | Herpes simplex virus-2† (% seropositive) | 7.3 | 6.4 | 15.1 | 15.7 | 16.1 | 12.7 |
| | Varicella zoster virus† (% seropositive) | 79.4 | 78.3 | 73.7 | 73 | 92.4 | 91.4 |
| | Epstein-Barr virus† (% seropositive) | 92.9 | 94.1 | 95.8 | 95.9 | 94.7 | 94.4 |
| | Cytomegalovirus† (% seropositive) | 54.2 | 51.8 | 79.9 | 80.1 | 58.2 | 52.4 |
| | Human herpesvirus-6A† (% seropositive) | 42.6 | 42.5 | 29.9 | 29.5 | 77.5 | 76.1 |
| | Human herpesvirus-6B† (% seropositive) | 53.9 | 50.9 | 43.7 | 44.2 | 79.2 | 77.1 |
| | Human betaherpesvirus-7† (% seropositive) | 72.7 | 74.0 | 47.2 | 46.3 | 94.7 | 94.2 |
| | Kaposi's sarcoma- | Low cell count | Low cell count | 2.0 | 1.9 | 8.1 | 8.0 |

| | | | | | | | |
|---|---|---|---|---|---|---|---|
| | associated virus† (% seropositive) | | | | | | |
| **Bacteria/Protozoa** | *C.trachomatis* (% seropositive) | 17.9 | 13.5 | 41.8 | 42.6 | 21.4 | 19.7 |
| | *H.pylori* (% seropositive) | 17.6 | 14.6 | 39.5 | 39.2 | 31.5 | 25.8 |
| | *T.gondii*† (% seropositive) | 24.4 | 23.5 | 21.2 | 20.8 | 28.0 | 25.9 |
| **Human Polyomaviruses** | BK virus (% seropositive) | 91.4 | 92.2 | 86.5 | 86.5 | 95.4 | 95.7 |
| | JC virus† (% seropositive) | 51.6 | 54.3 | 59.7 | 60.4 | 57.4 | 57.2 |
| | Merkel Cell virus (% seropositive) | 59.7 | 65.5 | 66.6 | 66.6 | 66.6 | 65.2 |
| **Human Papillomaviruses** | Human Papillomavirus-16 (% seropositive) | 2.7 | 2.7 | 4.8 | 4.8 | 4.4 | 4.8 |
| | Human Papillomavirus-18 (% seropositive) | 2.4 | 3.4 | 4.6 | 4.7 | 2.7 | 2.6 |
| **Pathogen burden index (mean (SD))** | | 0.43 (0.12) | 0.43 (0.12) | 0.47 (0.13) | 0.47 (0.13) | 0.53 (0.12) | 0.51 (0.12) |
| **Neurotropic pathogen burden index mean (SD))** | | 0.50 (0.15) | 0.49 (0.15) | 0.50 (0.14) | 0.50 (0.14) | 0.61 (0.15) | 0.59 (0.15) |
| **Ethnicity, N (%)** | | White* | White* | African Caribbean: 15.9 European: 47.8 South Asian: 36.3 Missing N: 4 | African Caribbean: 16.2 European: 46.7 South Asian: 37.1 | Asian or Asian British: 2.1 Black or Black British: 1.5 Chinese: 0.5 | Asian or Asian British: 1.4 Black or Black British: 0.7 Chinese: 0.4 |

Supplementary Tables

| | | | | | | | |
|---|---|---|---|---|---|---|---|
| | | | | | | Mixed: 0.5<br>Other: 0.8<br>White: 94.7<br>Missing N: 45 | Mixed: 0.4<br>Other: 0.4<br>White: 96.6<br>Missing N: 1 |
| **Body mass index in kg/m$^2$, mean (SD)** | | 27.7 (4.7)<br>Missing N: 5 | 27.5 (4.1) | 27.5 (4.7)<br>Missing N: 7 | 27.4 (4.5) | 27.3 (4.8)<br>Missing N: 32 | 26.5 (4.2) |
| **Alcohol intake, N (%)** | Low | 54.9 | 50.2 | 70.9 | 70.6 | 52 | 48.1 |
| | Moderate | 24.3 | 26.7 | 15.2 | 15.3 | 24.1 | 27 |
| | Heavy | 20.7<br>Missing N: 55 | 23<br>Missing N: 4 | 13.9<br>Missing N: 44 | 14.1<br>Missing N: 33 | 23.9<br>Missing N: 139 | 24.9<br>Missing N: 5 |
| **Ever smoker, N (%)** | | 53<br>Missing M: 42 | 45.9<br>Missing N: 7 | 43.3<br>Missing N: 14 | 42.6<br>Missing N: 8 | 44.2<br>Missing N: 54 | 37.5<br>Missing N: 2 |
| **Highest educational attainment, N (%)** | None | 36.2 | 20.4 | 31.1 | 30.1 | 17.2 | 7.3 |
| | Up to ordinary ('O') level, or equivalent | 20.8 | 24.6 | 20.1 | 20.1 | 27 | 21.6 |
| | Advanced ('A') level or equivalent, or higher | 43<br>Missing N: 89 | 54.9<br>Missing N: 12 | 48.8<br>Missing N: 239 | 49.9<br>Missing N: 212 | 55.8<br>Missing N: 105 | 71.1<br>Missing N: 1 |
| **Socioeconomic position, N (%)** | Professional | 7.9 | 12.6 | 3.5 | 3.7 | Townsend quintiles<br>Missing N: 8 | Townsend quintiles<br>Missing N: 1 |
| | Intermediate | 42 | 51.8 | 20.5 | 20.7 | | |
| | Nonmanual skilled | 23 | 20.8 | 15 | 14 | | |
| | Manual skilled | 14.6 | 9.1 | 35.3 | 35.3 | | |

| | | | | |
|---|---|---|---|---|
| Partly skilled or Unskilled‡ | 12.4<br>Missing<br>N: 11 | 5.7 | 25.4<br>Missing<br>N: 19 | 25.3<br>Missing<br>N: 14 |

Abbreviations: NSHD = National Survey of Health and Development; SABRE = Southall and Brent Revisited; UKB = UK Biobank. *The NSHD is predominantly white British as they are representative of births in mainland Britain in 1946, which was prior to major immigration flows [13]. † Pathogen considered to be neurotropic. ‡Categories collapsed due to low cell counts for the purpose of this table only

**Supplementary Table 2. Meta-analysed associations of serostatus and pathogen burden scores with brain volume.**
Estimates, 95% confidence intervals, p values, and $I^2$ statistics are presented. For pathogen burden analyses, estimates represent the difference in outcome per 0.1 unit increase in pathogen burden (proportion of pathogens or neurotropic pathogens seropositive to). Associations significant at p<0.05 are in bold, and those significant following multiple testing correction are further indicated by an asterisk. Model 1 included adjustments for total intracranial volume and other technical covariates; model 2 additionally for age, sex, and ethnicity; and model 3 additionally for BMI, smoking status, educational attainment, socioeconomic position, and alcohol intake.

| | **Brain volume** | | | | | | | | |
|---|---|---|---|---|---|---|---|---|---|
| | **Model 1** | | | **Model 2** | | | **Model 3** | | |
| | β [95% CI] (ml) | p | $I^2$ | β [95% CI (ml) | p | $I^2$ | β [95% CI] (ml) | p | $I^2$ |
| **HSV1** | 1.87 [-0.49,4.22] | 0.12 | 11 | 1.48 [-1.26,4.23] | 0.29 | 50 | 2.35 [-1.04,5.73] | 0.17 | 67 |
| **HSV2** | 4.33 [-5.18,13.85] | 0.37 | 88 | 2.87 [-5.81,11.54] | 0.52 | 89 | 2.73 [-4.73,10.19] | 0.47 | 85 |
| **VZV** | -0.58 [-2.95,1.79] | 0.63 | 0 | 0.03 [-2,2.07] | 0.97 | 0 | 0.04 [-1.97,2.06] | 0.97 | 0 |
| **EBV** | 3.07 [-4.74,10.88] | 0.44 | 64 | 1.21 [-5.19,7.62] | 0.71 | 61 | 2.45 [-2.98,7.88] | 0.38 | 46 |
| **CMV** | -0.56 [-4.56,3.43] | 0.78 | 69 | **2.57 [0.66,4.48]** | **0.008** | 0 | **2.62 [0.71,4.53]** | **0.007** | 0 |
| **HHV6A** | **-2.74 [-4.82,-0.66]** | **0.01** | 0 | -1.68 [-3.45,0.09] | 0.06 | 0 | -1.61 [-3.37,0.15] | 0.07 | 0 |
| **HHV6B** | 1.36 [-0.63,3.34] | 0.18 | 0 | 0.17 [-1.52,1.85] | 0.85 | 0 | 0 [-1.68,1.67] | 1 | 0 |
| **HHV7** | 2.24 [-0.79,5.26] | 0.15 | 26 | -0.46 [-3.62,2.71] | 0.78 | 41 | -0.56 [-3.99,2.86] | 0.75 | 49 |

Supplementary Tables

| | | | | | | | | | |
|---|---|---|---|---|---|---|---|---|---|
| **KSHV** | -2.4 [-7.75,2.94] | 0.38 | 0 | 0.02 [-4.26,4.31] | 0.99 | 0 | 0.07 [-4.21,4.35] | 0.97 | 0 |
| **HPV16** | 6.05 [-1.71,13.81] | 0.13 | 57 | 3.35 [-1.6,8.29] | 0.18 | 30 | 3.37 [-1.76,8.51] | 0.2 | 35 |
| **HPV18** | 3.58 [-1.31,8.46] | 0.15 | 0 | 2.9 [-1.27,7.07] | 0.17 | 0 | 3.18 [-0.97,7.32] | 0.13 | 0 |
| **BK** | 4.87 [-2.71,12.45] | 0.21 | 72 | 1.5 [-1.22,4.23] | 0.28 | 0 | 1.69 [-1.37,4.74] | 0.28 | 10 |
| **JC** | **-3.89 [-5.81,-1.97]** | **7E-05*** | 0 | -1.51 [-3.31,0.29] | 0.1 | 12 | -1.59 [-3.55,0.36] | 0.11 | 24 |
| **MCV** | -0.57 [-4.07,2.94] | 0.75 | 62 | -0.16 [-2.47,2.14] | 0.89 | 39 | 0 [-2.38,2.37] | 1 | 43 |
| **Ct** | 1.8 [-3.2,6.8] | 0.48 | 73 | -0.35 [-2.32,1.61] | 0.73 | 0 | -0.03 [-1.98,1.93] | 0.98 | 0 |
| **Tg** | -0.18 [-6.77,6.42] | 0.96 | 87 | 0.69 [-3.42,4.8] | 0.74 | 75 | 0.67 [-2.88,4.22] | 0.71 | 67 |
| **Hp** | -0.36 [-2.45,1.72] | 0.73 | 0 | 1.18 [-0.61,2.96] | 0.2 | 0 | 1.59 [-0.2,3.38] | 0.08 | 0 |
| **PBI** | 0.12 [-1.1,1.34] | 0.85 | 53 | 0.24 [-0.45,0.92] | 0.5 | 0 | 0.42 [-0.26,1.11] | 0.22 | 0 |
| **Neuro PBI** | -0.18 [-1.51,1.16] | 0.8 | 73 | 0.03 [-0.54,0.6] | 0.91 | 0 | 0.15 [-0.42,0.72] | 0.6 | 0 |

Abbreviations: BK=BK virus; CMV=Cytomegalovirus; Ct=*C.trachomatis*; EBV=Epstein-Barr virus; HHV=Human herpesvirus; Hp=*H.pylori*; HPV=Human Papillomavirus; HSV=Herpes simplex virus; JC=John Cunningham virus; KSHV=Kaposi's sarcoma-associated herpesviruses; MCV=Merkel Cell virus; PBI=Pathogen burden index; Tg=*T.gondii*; VZV=Varicella zoster virus. The neuro PBI included the nine herpesviruses, JC virus, and *T.gondii*.

**Supplementary Table 3. Meta-analysed associations of serostatus and pathogen burden scores with hippocampal volume.**
Estimates, 95% confidence intervals, p values, and $I^2$ statistics are presented. For pathogen burden analyses, estimates represent the difference in outcome per 0.1 unit increase in pathogen burden (proportion of pathogens or neurotropic pathogens seropositive to). Associations significant at p<0.05 are in bold, and those significant following multiple testing correction are further indicated by an asterisk. Model 1 included adjustments for total intracranial volume and other technical covariates; model 2 additionally for age, sex, and ethnicity; and model 3 additionally for BMI, smoking status, educational attainment, socioeconomic position, and alcohol intake.

| | **Hippocampal volume** | | | | | | | | |
|---|---|---|---|---|---|---|---|---|---|
| | **Model 1** | | | **Model 2** | | | **Model 3** | | |
| | β [95% CI] (ml) | p | $I^2$ | β [95% CI] (ml) | p | $I^2$ | β [95% CI] (ml) | p | $I^2$ |
| **HSV1** | 0 [-0.06,0.06] | 0.93 | 39 | 0.02 [-0.03,0.06] | 0.49 | 0 | 0.02 [-0.03,0.06] | 0.44 | 0 |
| **HSV2** | 0.01 [-0.1,0.13] | 0.81 | 63 | 0.06 [-0.01,0.12] | 0.07 | 0 | 0.06 [0,0.12] | 0.07 | 0 |
| **VZV** | **-0.06 [-0.11,0]** | **0.03** | 0 | **-0.07 [-0.12,-0.02]** | **0.009** | 0 | **-0.07 [-0.12,-0.02]** | **0.008** | 0 |
| **EBV** | 0.06 [-0.03,0.16] | 0.21 | 0 | 0.08 [-0.01,0.17] | 0.09 | 0 | 0.08 [-0.01,0.18] | 0.09 | 0 |
| **CMV** | -0.01 [-0.05,0.04] | 0.83 | 0 | 0.03 [-0.02,0.08] | 0.21 | 0 | 0.03 [-0.02,0.08] | 0.2 | 0 |
| **HHV6A** | -0.01 [-0.06,0.03] | 0.61 | 0 | 0.01 [-0.03,0.06] | 0.6 | 0 | 0.01 [-0.03,0.06] | 0.54 | 0 |
| **HHV6B** | -0.01 [-0.05,0.03] | 0.63 | 0 | -0.02 [-0.06,0.03] | 0.47 | 17 | -0.01 [-0.06,0.03] | 0.52 | 8 |
| **HHV7** | 0.01 [-0.04,0.07] | 0.61 | 11 | 0.02 [-0.03,0.06] | 0.53 | 0 | 0.01 [-0.04,0.06] | 0.61 | 0 |

| | | | | | | | | | |
|---|---|---|---|---|---|---|---|---|---|
| **KSHV** | **-0.12 [-0.23,-0.01]** | **0.04** | 0 | -0.08 [-0.19,0.02] | 0.12 | 0 | -0.08 [-0.19,0.02] | 0.13 | 0 |
| **HPV16** | 0.06 [-0.11,0.23] | 0.47 | 61 | 0.06 [-0.07,0.18] | 0.37 | 33 | 0.06 [-0.06,0.19] | 0.33 | 34 |
| **HPV18** | 0.02 [-0.1,0.15] | 0.71 | 17 | 0.02 [-0.09,0.13] | 0.68 | 4 | 0.04 [-0.09,0.17] | 0.56 | 27 |
| **BK** | 0.14 [-0.03,0.31] | 0.11 | 77 | 0.11 [-0.01,0.23] | 0.08 | 57 | 0.12 [-0.01,0.25]† | 0.08 | 62 |
| **JC** | **-0.06 [-0.1,-0.02]** | **0.007** | 0 | **-0.06 [-0.1,-0.02]** | **0.006** | 0 | **-0.05 [-0.09,-0.01]** | **0.01** | 0 |
| **MCV** | 0.03 [-0.04,0.1] | 0.38 | 55 | 0.03 [-0.03,0.1] | 0.31 | 52 | 0.03 [-0.04,0.1]† | 0.33 | 59 |
| **Ct** | -0.01 [-0.08,,0.06] | 0.73 | 41 | 0 [-0.05,0.05] | 0.97 | 0 | 0 [-0.05,0.05] | 0.86 | 0 |
| **Tg** | -0.03 [-0.08,0.02] | 0.2 | 0 | 0 [-0.05,0.05] | 0.99 | 0 | 0 [-0.05,0.04] | 0.84 | 0 |
| **Hp** | 0.03 [-0.03,0.09] | 0.26 | 33 | 0.04 [-0.01,0.09] | 0.1 | 10 | 0.03 [-0.01,0.08] | 0.13 | 0 |
| **PBI** | 0 [-0.03,0.02] | 0.71 | 41 | 0.01 [-0.01,0.02] | 0.43 | 0 | 0.01 [-0.01,0.02] | 0.44 | 0 |
| **Neuro PBI** | **-0.01 [-0.03,0]** | **0.05** | 0 | 0 [-0.02,0.01] | 0.65 | 0 | 0 [-0.02,0.01] | 0.66 | 0 |

Abbreviations: BK=BK virus; CMV=Cytomegalovirus; Ct=*C.trachomatis*; EBV=Epstein-Barr virus; HHV=Human herpesvirus; Hp=*H.pylori*; HPV=Human Papillomavirus; HSV=Herpes simplex virus; JC=John Cunningham virus; KSHV=Kaposi's sarcoma-associated herpesviruses; MCV=Merkel Cell virus; PBI=Pathogen burden index; Tg=*T.gondii*; VZV=Varicella zoster virus. The neuro PBI included the nine herpesviruses, JC virus, and *T.gondii*.

Supplementary Tables

**Supplementary Table 4. Meta-analysed associations of serostatus and pathogen burden scores with white matter lesion volume.**
Estimates, 95% confidence intervals, p values, and $I^2$ statistics are presented. For pathogen burden analyses, estimates represent the difference in outcome per 0.1 unit increase in pathogen burden (proportion of pathogens or neurotropic pathogens seropositive to). Associations significant at p<0.05 are in bold, and those significant following multiple testing correction are further indicated by an asterisk. Model 1 included adjustments for total intracranial volume and other technical covariates; model 2 additionally for age, sex, and ethnicity; and model 3 additionally for BMI, smoking status, educational attainment, socioeconomic position, and alcohol intake.

| | **White matter lesion volume** | | | | | | | | |
|---|---|---|---|---|---|---|---|---|---|
| | **Model 1** | | | **Model 2** | | | **Model 3** | | |
| | % change [95% CI] | p | $I^2$ | % change [95% CI] | p | $I^2$ | % change [95% CI] | p | $I^2$ |
| **HSV1** | 4.85 [-3.39,13.09] | 0.25 | 0 | 2.08 [-5.47,9.62] | 0.59 | 0 | 0.8 [-6.88,8.47] | 0.84 | 0 |
| **HSV2** | 13.59 [-4.51,31.68] | 0.14 | 55 | 2.36 [-8.02,12.74] | 0.66 | 0 | 0.94 [-9.47,11.35] | 0.86 | 0 |
| **VZV** | -5.26 [-14.52,3.99] | 0.26 | 0 | -4.47 [-13.63,4.7] | 0.34 | 8 | -3.82 [-14.6,6.96] | 0.49 | 25 |
| **EBV** | 7.03 [-9.89,23.94] | 0.42 | 0 | 5.03 [-10.45,20.51] | 0.52 | 0 | 2 [-13.66,17.66] | 0.8 | 0 |
| **CMV** | 11.77 [3.67,19.87] | **0.004** | 0 | 3.61 [-4.15,11.36] | 0.36 | 0 | 2.84 [-4.98,10.67] | 0.48 | 0 |
| **HHV6A** | 10.97 [3.03,18.92] | **0.007** | 0 | 5.5 [-1.77,12.77] | 0.14 | 0 | 5.48 [-1.81,12.76] | 0.14 | 0 |
| **HHV6B** | -0.57 [-8.17,7.03] | 0.88 | 0 | 2.06 [-4.86,8.97] | 0.56 | 0 | 2.79 [-4.17,9.75] | 0.43 | 0 |
| **HHV7** | -2.97 [-11.6,5.66] | 0.5 | 0 | 0.52 [-7.39,8.43] | 0.9 | 0 | 2.02 [-7.25,11.29] | 0.67 | 11 |

| | | | | | | | | | |
|---|---|---|---|---|---|---|---|---|---|
| **KSHV** | 26.72 [-3.35,56.79] | 0.08 | 37 | 14.61 [-7.55,36.77] | 0.2 | 19 | 16.14 [-8.48,40.77] | 0.2 | 28 |
| **HPV16** | 19.04 [1.75,36.32] | 0.03 | 0 | **20.48 [4.77,36.18]** | **0.01** | 0 | **20.78 [5.03,36.53]** | **0.01** | 0 |
| **HPV18** | 6.71 [-13.74,27.16] | 0.52 | 9 | 8.48 [-10.49,27.46] | 0.38 | 10 | 9.93 [-7.48,27.33] | 0.26 | 0 |
| **BK** | -20.19 [-55.61,15.23] | 0.26 | 82 | -11.77 [-30.99,7.45] | 0.23 | 49 | -10.03 [-30.32,10.26] † | 0.33 | 53 |
| **JC** | -2.26 [-18.74,14.22] | 0.79 | 78 | -3 [-15.5,9.5] | 0.64 | 66 | -3.64 [-16.18,8.9] | 0.57 | 67 |
| **MCV** | -6.12 [-22.35,10.12] | 0.46 | 75 | -7.42 [-21.24,6.4] | 0.29 | 70 | -6.58 [-20.09,6.93] | 0.34 | 69 |
| **Ct** | -4.5 [-25.19,16.18] | 0.67 | 78 | -3.39 [-15.33,8.54] | 0.58 | 43 | -4.07 [-17.81,9.67] | 0.56 | 55 |
| **Tg** | 10.23 [-1.74,22.21] | 0.09 | 44 | 3.67 [-4.19,11.52] | 0.36 | 0 | 3.04 [-4.84,10.92] | 0.45 | 0 |
| **Hp** | 6.86 [-1.06,14.79] | 0.09 | 0 | 2.76 [-4.58,10.09] | 0.46 | 0 | 1.1 [-6.33,8.52] | 0.77 | 0 |
| **PBI** | 2.88 [-3.54,9.29] | 0.38 | 76 | 0.96 [-3.42,5.34] | 0.67 | 53 | 0.82 [-3.81,5.44] | 0.73 | 57 |
| **Neuro PBI** | 3.86 [0.74,6.99] | **0.02** | 32 | 1.58 [-0.75,3.91] | 0.18 | 0 | 1.41 [-0.93,3.76] | 0.24 | 0 |

Abbreviations: BK=BK virus; CMV=Cytomegalovirus; Ct=*C.trachomatis*; EBV=Epstein-Barr virus; HHV=Human herpesvirus; Hp=*H.pylori*; HPV=Human Papillomavirus; HSV=Herpes simplex virus; JC=John Cunningham virus; KSHV=Kaposi's sarcoma-associated herpesviruses; MCV=Merkel Cell virus; PBI=Pathogen burden index; Tg=*T.gondii*; VZV=Varicella zoster virus. The neuro PBI included the nine herpesviruses, JC virus, and *T.gondii*.

**Supplementary Table 5. Meta-analysis results for APOE e2 and e4 and pathogen serostatus (and burden) interactions and neuroimaging outcomes.** P values are presented and full results are available on request. Associations significant at p<0.05 are in bold, and no associations survived multiple testing correction. Model 1 included adjustments for total intracranial volume and other technical covariates, and 10 genetic principal components; model 2 additionally for age and sex; and model 3 additionally for BMI, smoking status, educational attainment, socioeconomic position, and alcohol intake.

| | | **Brain volume** | | | **Hippocampal volume** | | | **White matter lesion volume** | | |
|---|---|---|---|---|---|---|---|---|---|---|
| | | Model 1 | Model 2 | Model 3 | Model 1 | Model 2 | Model 3 | Model 1 | Model 2 | Model 3 |
| | | p | p | p | p | p | p | p | p | p |
| ***Serostatus*** | | | | | | | | | | |
| **HSV1†*** | *APOE* e2 | 0.94 | 0.74 | 0.82 | 0.34 | 0.35 | 0.29 | 0.87 | 0.71 | 0.72 |
| | *APOE* e4 | 0.28 | 0.47 | 0.46 | 0.25 | 0.34 | 0.38 | 0.64 | 0.48 | 0.44 |
| **HSV2†** | *APOE* e2 | 0.61 | 0.45 | 0.39 | 0.71 | 0.77 | 0.79 | 0.26 | 0.3 | 0.27 |
| | *APOE* e4 | 0.62 | 0.81 | 0.74 | 0.73 | 0.63 | 0.6 | 0.17 | 0.08 | 0.09 |
| **VZV†** | *APOE* e2 | 0.98 | 0.87 | 0.7 | 0.29 | 0.28 | 0.25 | 0.93 | 0.9 | 0.67 |
| | *APOE* e4 | 0.94 | 0.96 | 0.98 | 0.58 | 0.62 | 0.58 | 0.93 | 0.73 | 0.59 |
| **EBV†** | *APOE* e2 | 0.65 | 0.71 | 0.85 | 0.38 | 0.64 | 0.38 | 0.16 | 0.55 | 0.59 |
| | *APOE* e4 | 0.69 | 0.99 | 0.95 | 0.47 | 0.72 | 0.83 | 0.76 | 0.68 | 0.61 |
| **CMV†*** | *APOE* e2 | 0.82 | 0.35 | 0.34 | 0.99 | 0.92 | 0.8 | 0.25 | 0.14 | 0.15 |
| | *APOE* e4 | 0.16 | 0.41 | 0.26 | 0.79 | 0.98 | 0.96 | 0.4 | 0.37 | 0.38 |
| **HHV6A†** | *APOE* e2 | 0.83 | 0.98 | 0.98 | 0.74 | 0.83 | 0.71 | 0.34 | 0.4 | 0.5 |
| | *APOE* e4 | 0.42 | 0.87 | 1 | 0.62 | 1 | 0.9 | 0.12 | 0.6 | 0.53 |
| **HHV6B†** | *APOE* e2 | 0.69 | 0.62 | 0.68 | 0.45 | 0.34 | 0.35 | 0.65 | 0.76 | 0.74 |
| | *APOE* e4 | 0.83 | 0.38 | 0.47 | 0.9 | 0.96 | 0.81 | 0.37 | 0.54 | 0.81 |
| **HHV7†** | *APOE* e2 | 0.69 | 0.37 | 0.52 | 0.58 | 0.52 | 0.52 | 0.84 | 0.63 | 0.63 |
| | *APOE* e4 | 0.47 | 0.84 | 0.47 | 0.52 | 0.33 | 0.35 | 0.96 | 0.74 | 0.77 |
| **KSHV†*** | *APOE* e2 | 0.55 | 0.71 | 0.87 | 0.33 | 0.34 | 0.36 | 0.73 | 0.86 | 0.73 |
| | *APOE* e4 | 0.76 | 0.59 | 0.44 | 0.37 | 0.63 | 0.6 | 0.54 | 0.61 | 0.5 |
| **HPV16*** | *APOE* e2 | 0.07 | 0.75 | 0.97 | 0.21 | 0.5 | 0.41 | 0.15 | 0.45 | 0.55 |
| | *APOE* e4 | 0.8 | 0.54 | 0.29 | 0.81 | 0.81 | 0.75 | 0.33 | 0.59 | 0.44 |
| **HPV18*** | *APOE* e2 | 0.76 | 0.35 | 0.45 | 0.38 | 0.25 | 0.27 | 0.28 | 0.26 | 0.28 |
| | *APOE* e4 | 0.36 | 0.49 | 0.23 | 0.32 | 0.32 | 0.21 | 0.59 | 0.65 | 0.72 |
| **BK** | *APOE* e2 | 0.79 | 0.98 | 0.75 | 0.74 | 0.84 | 0.68 | 0.45 | 0.46 | 0.56 |
| | *APOE* e4 | 0.63 | 0.63 | 0.29 | 0.93 | 0.9 | 0.97 | 0.88 | 0.59 | 0.68 |

| | | | | | | | | | | |
|---|---|---|---|---|---|---|---|---|---|---|
| **JC†** | *APOE* e2 | 0.44 | 0.72 | 0.91 | 0.19 | 0.25 | 0.26 | 0.29 | 0.44 | 0.37 |
| | *APOE* e4 | 0.85 | 0.6 | 0.6 | 0.45 | 0.24 | 0.22 | 0.52 | 0.3 | 0.25 |
| **MCV** | *APOE* e2 | 0.31 | 0.61 | 0.56 | 0.09 | 0.08 | **0.04** | 0.06 | 0.18 | 0.17 |
| | *APOE* e4 | 0.5 | 0.34 | 0.28 | 0.79 | 0.6 | 0.51 | 0.39 | 0.55 | 0.55 |
| **Ct** | *APOE* e2 | 0.99 | 0.98 | 0.75 | 0.38 | 0.34 | 0.31 | 0.32 | 0.26 | 0.28 |
| | *APOE* e4 | 0.35 | 0.37 | 0.23 | 0.76 | 0.53 | 0.58 | 0.88 | 0.82 | 0.74 |
| **Tg†** | *APOE* e2 | 0.42 | 0.93 | 0.85 | **0.003** | **0.004** | **0.004** | 0.14 | 0.13 | 0.1 |
| | *APOE* e4 | 0.86 | 0.49 | 0.38 | 0.44 | 0.28 | 0.27 | 0.94 | 0.69 | 0.83 |
| **Hp** | *APOE* e2 | 0.76 | 0.94 | 0.77 | 0.29 | 0.23 | 0.21 | 0.36 | 0.45 | 0.44 |
| | *APOE* e4 | 0.27 | 0.47 | 0.49 | 0.96 | 0.87 | 0.71 | 0.59 | 0.41 | 0.3 |
| ***Pathogen burden*** | | | | | | | | | | |
| **PBI** | *APOE* e2 | 0.56 | 0.66 | 0.62 | **0.02** | **0.01** | **0.004** | 0.07 | 0.07 | 0.07 |
| | *APOE* e4 | 0.71 | 0.72 | 0.74 | 0.81 | 0.69 | 0.62 | 0.75 | 0.91 | 0.94 |
| **Neurotropic PBI** | *APOE* e2 | 0.44 | 0.49 | 0.54 | **0.03** | **0.03** | **0.01** | 0.13 | 0.1 | 0.1 |
| | *APOE* e4 | 0.5 | 0.93 | 0.98 | 0.82 | 0.58 | 0.65 | 0.76 | 0.93 | 0.8 |

Abbreviations: BK=BK virus; CMV=Cytomegalovirus; Ct=*C.trachomatis*; EBV=Epstein-Barr virus; HHV=Human herpesvirus; Hp=*H.pylori*; HPV=Human papillomavirus; HSV=Herpes-simplex virus; JC=John Cunningham virus; KSHV=Kaposi's sarcoma-associated herpesviruses; MCV=Merkel Cell virus; PBI=Pathogen burden index; Tg=*T.gondii*; VZV=Varicella zoster virus. †= pathogen considered to be neurotropic .*analyses did not run in all studies due to insufficient degrees of freedom.

**Supplementary Table 6. Meta-analysis results for associations of Merkel Cell virus, *T.gondii*, and pathogen burden indices with hippocampal volume, stratified by APOE e2 carrier status.**

Estimates, 95% confidence intervals, and p values are presented. For pathogen burden exposures, estimates represent the change in outcome per 0.1 unit increase in burden index. Associations significant at p<0.05 are in bold. Model 1 included adjustments for total intracranial volume and other technical covariates; model 2 additionally for age and sex; and model 3 additionally for BMI, smoking status, educational attainment, socioeconomic position, and alcohol intake.

| | | Hippocampal volume | | | | | |
|---|---|---|---|---|---|---|---|
| | | Model 1 | | Model 2 | | Model 3 | |
| | | β[95%CI] (ml) | p | β[95%CI] (ml) | p | β[95%CI] (ml) | p |
| **Merkel Cell virus** | *APOE* e2 carrier | -0.2 [-0.43,0.02] | 0.08 | -0.17 [-0.41,0.06] | 0.15 | -0.23 [-0.47,0.01] | 0.06 |
| | *APOE* e2 non-carrier | 0.05 [0,0.11] | 0.06 | 0.05 [0,0.1] | 0.07 | 0.05 [0,0.1] | 0.07 |
| ***T.gondii*** | *APOE* e2 carrier | **-0.22 [-0.36,-0.07]** | **0.003** | **-0.18 [-0.32,-0.03]** | **0.02** | -0.15 [-0.32,0.02] | 0.08 |
| | *APOE* e2 non-carrier | -0.03 [-0.09,0.03] | 0.38 | 0 [-0.06,0.06] | 0.99 | -0.01 [-0.07,0.05] | 0.81 |
| **Pathogen burden index** | *APOE* e2 carrier | **-0.09 [-0.15,-0.02]** | **0.01** | **-0.07 [-0.13,-0.02]** | **0.009** | **-0.07 [-0.13,-0.01]** | **0.01** |
| | *APOE* e2 non-carrier | 0 [-0.02,0.03] | 0.69 | 0.01 [-0.01,0.03] | 0.41 | 0.01 [-0.01,0.03] | 0.45 |
| **Neurotropic pathogen burden index** | *APOE* e2 carrier | **-0.05 [-0.09,-0.01]** | **0.01** | **-0.05 [-0.09,0]** | **0.03** | -0.04 [-0.09,0] | 0.06 |
| | *APOE* e2 non-carrier | -0.01 [-0.03,0.01] | 0.32 | 0 [-0.02,0.01] | 0.74 | 0 [-0.02,0.02] | 0.79 |

**Supplementary Table 7. Meta-analysed associations of seroreactivity tertiles with brain volume.**
Estimates, 95% confidence intervals, p values, and $I^2$ statistics are presented. Associations significant at $p<0.05$ are in bold, and those significant following multiple testing correction are further indicated by an asterisk. Model 1 included adjustments for total intracranial volume and other technical covariates; model 2 additionally for age, sex, and ethnicity; and model 3 additionally for BMI, smoking status, educational attainment, socioeconomic position, and alcohol intake.

| | **Brain volume** | | | | | | | | |
|---|---|---|---|---|---|---|---|---|---|
| | **Model 1** | | | **Model 2** | | | **Model 3** | | |
| | β [95% CI] | p | $I^2$ | β [95% CI] | p | $I^2$ | β [95% CI] | p | $I^2$ |
| **HSV1** | -0.37 [-2.33,1.6] | 0.71 | 0 | 0.12 [-1.58,1.81] | 0.89 | 0 | 0.18 [-1.52,1.87] | 0.84 | 0 |
| **HSV2** | -0.85 [-5.95,4.25] | 0.74 | 0 | -1.58 [-5.7,2.54] | 0.45 | 0 | -0.85 [-5.1,3.41] | 0.7 | 0 |
| **VZV** | -1.43 [-3.3,0.43] | 0.13 | 0 | -1.1 [-2.67,0.47] | 0.17 | 0 | -1.18 [-2.74,0.39] | 0.14 | 0 |
| **EBV** | -1.09 [-5.25,3.07] | 0.61 | 81 | -0.41 [-1.84,1.03] | 0.58 | 0 | -0.31 [-1.73,1.12] | 0.67 | 0 |
| **CMV** | -0.32 [-4.23,3.6] | 0.87 | 67 | -0.62 [-2.5,1.25] | 0.52 | 12 | -0.55 [-2.23,1.12] | 0.52 | 0 |
| **HHV6A** | 1.37 [-2.16,4.9] | 0.45 | 42 | 0.71 [-1.42,2.83] | 0.52 | 0 | 0.77 [-1.68,3.21] | 0.54 | 18 |
| **HHV6B** | 1.32 [-1,3.65] | 0.26 | 0 | -1.02 [-2.93,0.88] | 0.29 | 0 | -1.12 [-3.01,0.78] | 0.25 | 0 |
| **HHV7** | -1.17 [-3.24,0.91] | 0.27 | 0 | -1.62 [-3.35,0.12] | 0.07 | 0 | -1.65 [-3.38,0.07] | 0.06 | 0 |
| **BK** | 0.92 [-0.78,2.63] | 0.29 | 0 | -0.45 [-1.9,1] | 0.54 | 0 | -0.59 [-2.04,0.85] | 0.42 | 0 |

Supplementary Tables

| | | | | | | | | | |
|---|---|---|---|---|---|---|---|---|---|
| **JC** | -1.47 [-4.77,1.82] | 0.38 | 47 | -0.37 [-3.37,2.63] | 0.81 | 54 | -0.63 [-4.33,3.07] | 0.74 | 69 |
| **MCV** | -3.54 [-7.71,0.64] | 0.1 | 73 | -3.52 [-8.45,1.41] | 0.16 | 86 | -3.02 [-7.93,1.88] | 0.23 | 85 |
| **Ct** | 1.05 [-5.5,7.59] | 0.75 | 63 | -0.14 [-4.81,4.52] | 0.95 | 46 | 1.2 [-4.64,7.04] | 0.69 | 60 |
| **Tg** | -4.46 [-9.68,0.77] | 0.09 | 0 | **-6.63 [-11.26,-2]** | **0.005** | 12 | **-7.11 [-14.13,-0.08]** | **0.05** | 50 |
| **Hp** | 2.89 [-0.21,5.99] | 0.07 | 0 | 1.86 [-0.84,4.55] | 0.18 | 0 | 1.84 [-0.9,4.59] | 0.19 | 0 |

Abbreviations: BK=BK virus; CMV=Cytomegalovirus; Ct=*C.trachomatis*; EBV=Epstein-Barr virus; HHV=Human herpesvirus; Hp=*H.pylori*; HSV=Herpes simplex virus; JC=John Cunningham virus; MCV=Merkel Cell virus; Tg=*T.gondii*; VZV=Varicella zoster virus. Kaposi's sarcoma-associated herpesviruses and Human Papillomavirus 16 and 18 were not tested as seroprevalence were <5% in at least one cohort.

**Supplementary Table 8. Meta-analysed associations of seroreactivity tertiles with hippocampal volume.**
Estimates, 95% confidence intervals, p values, and $I^2$ statistics are presented. Associations significant at $p<0.05$ are in bold, and those significant following multiple testing correction are further indicated by an asterisk. Model 1 included adjustments for total intracranial volume and other technical covariates; model 2 additionally for age, sex, and ethnicity; and model 3 additionally for BMI, smoking status, educational attainment, socioeconomic position, and alcohol intake.

| | Hippocampal volume | | | | | | | | |
|---|---|---|---|---|---|---|---|---|---|
| | Model 1 | | | Model 2 | | | Model 3 | | |
| | β [95% CI] | p | $I^2$ | β [95% CI] | p | $I^2$ | β [95% CI] | p | $I^2$ |
| **HSV1** | 0 [-0.05,0.04] | 0.9 | 0 | -0.02 [-0.07,0.02] | 0.28 | 0 | -0.02 [-0.07,0.02] | 0.28 | 0 |
| **HSV2** | 0 [-0.1,0.11] | 0.96 | 7 | 0.02 [-0.1,0.14] | 0.73 | 14 | 0.01 [-0.15,0.16] | 0.92 | 34 |
| **VZV** | -0.05 [-0.11,0.02] | 0.18 | 62 | -0.05 [-0.11,0] | 0.063 | 45 | -0.05 [-0.11,0.01] | 0.07 | 47 |
| **EBV** | -0.02 [-0.06,0.01] | 0.19 | 0 | 0 [-0.04,0.03] | 0.94 | 0 | 0 [-0.04,0.03] | 0.87 | 0 |
| **CMV** | -0.02 [-0.06,0.03] | 0.44 | 0 | -0.01 [-0.05,0.04] | 0.78 | 0 | -0.01 [-0.05,0.03] | 0.72 | 0 |
| **HHV6A** | 0.01 [-0.05,0.06] | 0.81 | 0 | 0.01 [-0.05,0.06] | 0.83 | 0 | 0 [-0.05,0.06] | 0.88 | 0 |
| **HHV6B** | **0.06 [0.01,0.11]** | **0.02** | 2 | 0.04 [0,0.09] | 0.07 | 0 | **0.05 [0,0.1]** | **0.04** | 0 |
| **HHV7** | -0.04 [-0.09,0] | 0.06 | 0 | -0.04 [-0.08,0.01] | 0.09 | 0 | -0.04 [-0.08,0.01] | 0.09 | 0 |
| **BK** | 0 [-0.04,0.04] | 0.99 | 2 | 0 [-0.04,0.03] | 0.95 | 0 | 0 [-0.04,0.03] | 0.87 | 0 |

| | | | | | | | | | |
|---|---|---|---|---|---|---|---|---|---|
| **JC** | -0.02 [-0.06,0.03] | 0.51 | 0 | -0.02 [-0.07,0.02] | 0.36 | 0 | -0.02 [-0.06,0.03] | 0.45 | 0 |
| **MCV** | -0.03 [-0.07,0.02] | 0.23 | 0 | -0.01 [-0.06,0.03] | 0.49 | 0 | -0.02 [-0.06,0.03] | 0.45 | 0 |
| **Ct** | 0.05 [-0.02,0.11] | 0.17 | 6 | **0.07 [0.01,0.13]** | **0.03** | 0 | **0.07 [0.01,0.14]** | **0.02** | 0 |
| **Tg** | 0.06 [-0.09,0.21] | 0.44 | 37 | 0.03 [-0.09,0.15] | 0.59 | 6 | 0 [-0.14,0.13] | 0.96 | 5 |
| **Hp** | 0.05 [-0.03,0.12] | 0.22 | 0 | 0.03 [-0.04,0.11] | 0.36 | 0 | 0.03 [-0.05,0.1] | 0.51 | 0 |

Abbreviations: BK=BK virus; CMV=Cytomegalovirus; Ct=*C.trachomatis*; EBV=Epstein-Barr virus; HHV=Human herpesvirus; Hp=*H.pylori*; HSV=Herpes simplex virus; JC=John Cunningham virus; MCV=Merkel Cell virus; Tg=*T.gondii*; VZV=Varicella zoster virus. Kaposi's sarcoma-associated herpesviruses and Human Papillomavirus 16 and 18 were not tested as seroprevalence were <5% in at least one cohort.

**Supplementary Table 9. Meta-analysed associations of seroreactivity tertiles with white matter lesion volume.**
Estimates, 95% confidence intervals, p values, and $I^2$ statistics are presented. Associations significant at $p<0.05$ are in bold, and those significant following multiple testing correction are further indicated by an asterisk. Model 1 included adjustments for total intracranial volume and other technical covariates; model 2 additionally for age, sex, and ethnicity; and model 3 additionally for BMI, smoking status, educational attainment, socioeconomic position, and alcohol intake.

| | White matter lesion volume | | | | | | | | |
|---|---|---|---|---|---|---|---|---|---|
| | Model 1 | | | Model 2 | | | Model 3 | | |
| | % change [95%CI] | p | $I^2$ | % change [95%CI] | p | $I^2$ | % change [95%CI] | p | $I^2$ |
| **HSV1** | -10.61 [-19.81,-1.41] | 0.02 | 27 | -7.19 [-13.99,-0.39] | **0.04** | 0 | -6.01 [-12.83,0.82] | 0.08 | 0 |
| **HSV2** | -1.9 [-21.52,17.73] | 0.85 | 0 | -0.41 [-18.24,17.43] | 0.96 | 0 | 0.56 [-18.21,19.32] | 0.95 | 0 |
| **VZV** | 2.48 [-7.71,12.67] | 0.63 | 48 | 3.7 [-3.67,11.07] | 0.32 | 19 | 4.53 [-5.93,14.99] | 0.4 | 56 |
| **EBV** | 12.12 [5.67,18.57] | **2e-4** | 0 | 4.58 [-1.36,10.52] | 0.13 | 0 | 4.44 [-1.53,10.4] | 0.15 | 0 |
| **CMV** | 8.08 [0.37,15.79] | **0.04** | 0 | 4.46 [-2.75,11.66] | 0.23 | 0 | 5.05 [-2.21,12.3] | 0.17 | 0 |
| **HHV6A** | 0.14 [-9.57,9.84] | 0.98 | 0 | 1.82 [-7.02,10.66] | 0.69 | 0 | 1.99 [-6.99,10.98] | 0.66 | 0 |
| **HHV6B** | -4 [-12.84,4.84] | 0.38 | 5 | 0.33 [-7.7,8.36] | 0.94 | 4 | 0.18 [-7.71,8.07] | 0.96 | 0 |
| **HHV7** | 7.44 [-0.12,15.01] | 0.05 | 0 | 5.52 [-1.46,12.49] | 0.12 | 0 | 5.37 [-1.65,12.39] | 0.13 | 0 |

Supplementary Tables

| | | | | | | | | | |
|---|---|---|---|---|---|---|---|---|---|
| **BK** | -5.93 [-12.38,0.51] | 0.07 | 0 | -5.67 [-11.61,0.28] | 0.06 | 0 | -6.1 [-12.08,-0.11] | **0.05** | 0 |
| **JC** | 1.53 [-6.9,9.96] | 0.72 | 7 | 3.78 [-3.76,11.31] | 0.33 | 3 | 1.95 [-9.23,13.13] | 0.73 | 49 |
| **MCV** | 5.15 [-2.48,12.78] | 0.19 | 0 | 1.56 [-5.41,8.54] | 0.66 | 0 | 1.52 [-5.52,8.55] | 0.67 | 0 |
| **Ct** | 7.12 [-11.39,25.63] | 0.45 | 41 | 5.33 [-5.33,15.99] | 0.33 | 0 | 6.87 [-4.04,17.78] | 0.22 | 0 |
| **Tg** | 6.71 [-17.88,31.31] | 0.59 | 36 | 5.58 [-11.46,22.61] | 0.52 | 0 | 4.63 [-13.49,22.75] | 0.62 | 0 |
| **Hp** | 14.01 [-25.93,53.95] | 0.49 | 80 | 14.71 [-23.7,53.12] | 0.45 | 81 | 9.36 [-26.28,44.99] | 0.61 | 76 |

Abbreviations: BK=BK virus; CMV=Cytomegalovirus; Ct=*C.trachomatis*; EBV=Epstein-Barr virus; HHV=Human herpesvirus; Hp=*H.pylori*; HSV=Herpes simplex virus; JC=John Cunningham virus; MCV=Merkel Cell virus; Tg=*T.gondii*; VZV=Varicella zoster virus. Kaposi's sarcoma-associated herpesviruses and Human Papillomavirus 16 and 18 were not tested as seroprevalence were <5% in at least one cohort.
