## Supplementary Notes for "Common infections and neuroimaging markers of dementia in three UK cohort studies"

### Serology measures

We used serostatus variables provided by the UK Biobank (UKB) [1], and derived serostatus in the MRC National Survey of Health and Development (NSHD) and Southall and Brent Revisited (SABRE) using cut-offs supplied by the developers of the multiplex serology platform (German Cancer Research Center in Heidelberg) [1,2]. UKB variables can be extracted using field codes, which are included below, alongside details of serostatus derivation NSHD and SABRE. In addition to the pathogens listed in the table below, antigen seroreactivity values for immunodeficiency virus-1, hepatitis B, hepatitis C, and human T-cell leukemia virus-1 were additionally assayed in UKB, and antigen seroreactivity values for *F.nucleatum* in NSHD and SABRE. As analyses were restricted to the 17 pathogens common to all three studies, these pathogens were not investigated.

| | UKB field codes | NSHD and SABRE |
|---|---|---|
| Herpes simplex virus-1 | 23050 | 1gG (170) |
| Herpes simplex virus-2 | 23051 | 2mgG unique (180) |
| Varicella zoster virus | 23052 | gE / gI (100) |
| Epstein-Barr virus | 23053 | ≥2 positive out of EBNA (411), EA-D (110), VCAp18 (2526), Zebra (74) |
| Cytomegalovirus | 23054 | ≥2 positive out of pp150 (100), pp52 (854), pp28 (100) |
| Human herpesvirus-6A | 23056 | IE1A (100) AND/OR p100 (75) |
| Human herpesvirus-6B | 23057 | IE1B (100) AND/OR p101K (100) |
| Human betaherpesvirus 7 | 23058 | U14 (225) |
| Kaposi's sarcoma-associated virus | 23059 | LANA 3 (100) AND K8.1 (100) |
| BK virus | 23065 | VP1 (150) |
| JC virus | 23066 | VP1 (150) |
| Merkel Cell virus | 23067 | VP1 (150) |
| Human Papillomavirus-16 | 23068 and 23075 | L1 (100) |
| Human Papillomavirus-18 | 23069 | L1 (100) |
| *T.gondii* | 23062 | p22 (150) AND/OR sag1 (150) |

| | | |
|---|---|---|
| *H.pylori* | 23073 and 23074 | ≥3 positive out of 8:<br>HP 10 (GroEL) (100)<br>HP1098 (Hcp C) (200)<br>HP 1564 (400)<br>HP 887/2 (VacA-C) (250)<br>HP547/1 (CagA-N) (400)<br>HP305 (150)<br>HP73 (UreA) (250)<br>HP875 (Catalase) (400) |
| *C.trachomatis* | 23070 and 23071 | pGP3 (95) |

In addition, in some cases, there were several antigens recommended by the developers of the multiplex serology platform for use in seroreactivity analyses - these are displayed below. In situations where there were multiple recommended antigens, we randomly selected one using the sample function in base R and followed up the others in sensitivity analyses (see table below).

| **Infectious agent group** | **Pathogen** | **N Antigens available** | **Recommended antigen** | |
|---|---|---|---|---|
| | | | **UKB** | **NSHD/SABRE** |
| Herpesviruses | Herpes simplex virus-1 | 1 | 1gG | 1gG |
| | Herpes simplex virus-2 | 1 | 2mgG unique | 2mgG unique |
| | Varicella zoster virus | 1 | gE/gI | gE/gI |
| | Epstein Barr virus | 4 | VCA p18 or EBNA | VCA p18 or EBNA |
| | Cytomegalovirus | 3 | pp150 or pp52 or pp28 | p150 or pp52 or pp28 |
| | Human herpesvirus-6A | 1-2 | 1E1A | 1E1A |

| | | | | |
|---|---|---|---|---|
| | Human herpesvirus-6B | 2 | 1E1B | 1E1B |
| | Human betaherpesvirus 7 | 1 | U14 | U14 |
| | Kaposi's sarcoma-associated virus | 2 | K8.1 | K8.1 |
| Polyomaviruses | BK virus | 1 | BK VP1 | BK VP1 |
| | JC virus | 1 | JC VP1 | JC VP1 |
| | Merkel Cell virus | 1 | MCV VP1 | MCV VP1 |
| Human papillomavirus | Human papillomavirus-16 | 3 | L1 | L1 |
| | Human papillomavirus-18 | 1 | L1 | L1 |
| Bacteria/Protozoa | *T.gondii* | 2 | p22 or sag-1 | p22 or sag-1 |
| | *H.pylori* | 6-8 | CagA-N | HP 10 (GroEL)<br>HP1098 (Hcp C)<br>HP 1564<br>HP 887/2 (VacA-C)<br>HP547/1 (CagA-N)<br>HP305<br>HP73 (UreA)<br>HP875 (Catalase) |
| | *C.trachomatis* | 1 | pGP3 | pGP3 |

**Genetic data quality control**

We used directly genotyped data for genetic quality control and to derive genetic principal components. Details of genotyping can be found for NSHD [3], SABRE [4], and UKB [5] elsewhere. Briefly, blood samples were collected at the age 53 visit for NSHD and baseline (visit 1) for SABRE, and participants were genotyped using the DrugDev array. Two genotyping arrays (BiLEVE Axiom and Affymetrix UK Biobank Axiom) were used to genotype UKB participants. NSHD and SABRE genetic QC were performed in house, and we used UKB provided QC criteria and genetic principal components (following their own central QC) for UKB. For

SABRE, QC were applied separately by self-reported ethnicity. Quality control (QC) for SABRE and NSHD included filtering for common (MAF>0.01) biallelic autosomal variants that did not exhibit significant missingness (call rate >98%) or deviate from Hardy-Weinberg equilibrium ($p<1e^{-4}$). Samples that did not exhibit significant heterozygosity (mean ±3sd) or missingness (call rate >98%), with concordant genetic and self-reported sex, and that were unrelated (KING >0.088) and closely clustered with reference panel populations (defined visually using genetic principal components) were additionally retained. Genetic principal components were generated in each study population following pruning and removal of variants in long range LD regions [6]. QC steps were performed using plink1.9 and plink2 [7], and principal components using smartpca in EIGENSOFT [8].

Similar QC steps were performed and supplied by the UKB [5]. Unrelated (KING>0.088) participants were retained that did not exhibit significant heterozygosity or missingness (field id: 22003), with concordant genetic and self-reported sex (field ids: 31 and 22001), that did not have sex chromosome aneuploidies (field id: 22019), that self-identified as "White-British" and were not PCA outliers (field id: 21000). UKB provided principal components which we used (field id: 22009).

**Imaging protocols**

A table indicating imaging acquisition protocols is included below.

| **Study** | **Scanner and field strength** | **T1** | | | | | **FLAIR** | | | |
|---|---|---|---|---|---|---|---|---|---|---|
| | | **Dura-tion (min:s)** | **Type** | **Voxel res** | **Matrix size** | **TI/TE/TR** | **Dura-tion (min:s)** | **Voxel res (mm)** | **Matrix** | **TI/TE/TR** |
| NSHD | PET/MR 3T | 5:06 | MPRAGE | 1.1x<br>1.1x<br>1.1 | 256x<br>256x<br>208 | TI-870<br>TE=2.92<br>TR=2000 | 6:27 | 1.1x<br>1.1x<br>1.1 | 256x<br>256x<br>176 | TI=1800<br>TE=402<br>TR=5000 |
| SABRE* | Signa HDx 1.5T | 3:32 | Spin Echo | 0.94x<br>0.94x<br>5 | 256x<br>256x<br>28 | TI=NA<br>TE=12<br>TR=380 | 7:12 | 0.47x<br>0.47x<br>3 | 512x<br>512x<br>48 | TI=2250<br>TE=148.3<br>TR=9002 |
| SABRE* | GE DISCOVERY MR750 3T | 4:12 | Spin Echo | 0.47x<br>0.47x<br>5 | 512x<br>512x<br>30 | TI=NA<br>TE=9<br>TR=420 | 7:19 | 0.47x<br>0.47x<br>3 | 512x<br>512x<br>50 | TI=2250<br>TE=149<br>TR=8775 |

| | | | | | | | | | | |
|---|---|---|---|---|---|---|---|---|---|---|
| UKB | | 4:54 | MPRAGE | 1x 1x 1 | 208x 208x 256 | TI= 880 TR= 2000 | 5:52 | 1.05x 1x 1 | 192x 256x 256 | TI=1800 TR=5000 |

* T1 SABRE scans were resampled to 1mm isotropic

**Covariates**

Covariates included total intracranial volume (derived in house), and other technical variables related to neuroimaging or serology data collection (NSHD: blood clinic, SABRE: scanner; UKB: scanner co-ordinates and blood clinic), age at serology and neuroimaging (if not collected at the same time point), sex, ethnicity (SABRE: African-Caribbean, European, South Asian; UKB: Asian or Asian British, Black or Black British, Chinese, Mixed, Other, White), highest educational qualification (none; up to ordinary ('O') level or equivalent; advanced ('A') level or equivalent, or higher), socioeconomic position (NSHD and SABRE: current or last known occupation categorised into six groups according to the UK Registrar General; UKB: Townsend deprivation index quintiles), BMI (kg/m2), smoking status (ever/never), and alcohol intake (low <7 units per week, medium 7-14 units per week, high >14 units per week). A directed acyclic graph can be found below, displaying our hypothesised relationships for covariates and confounders of serology-outcome associations.

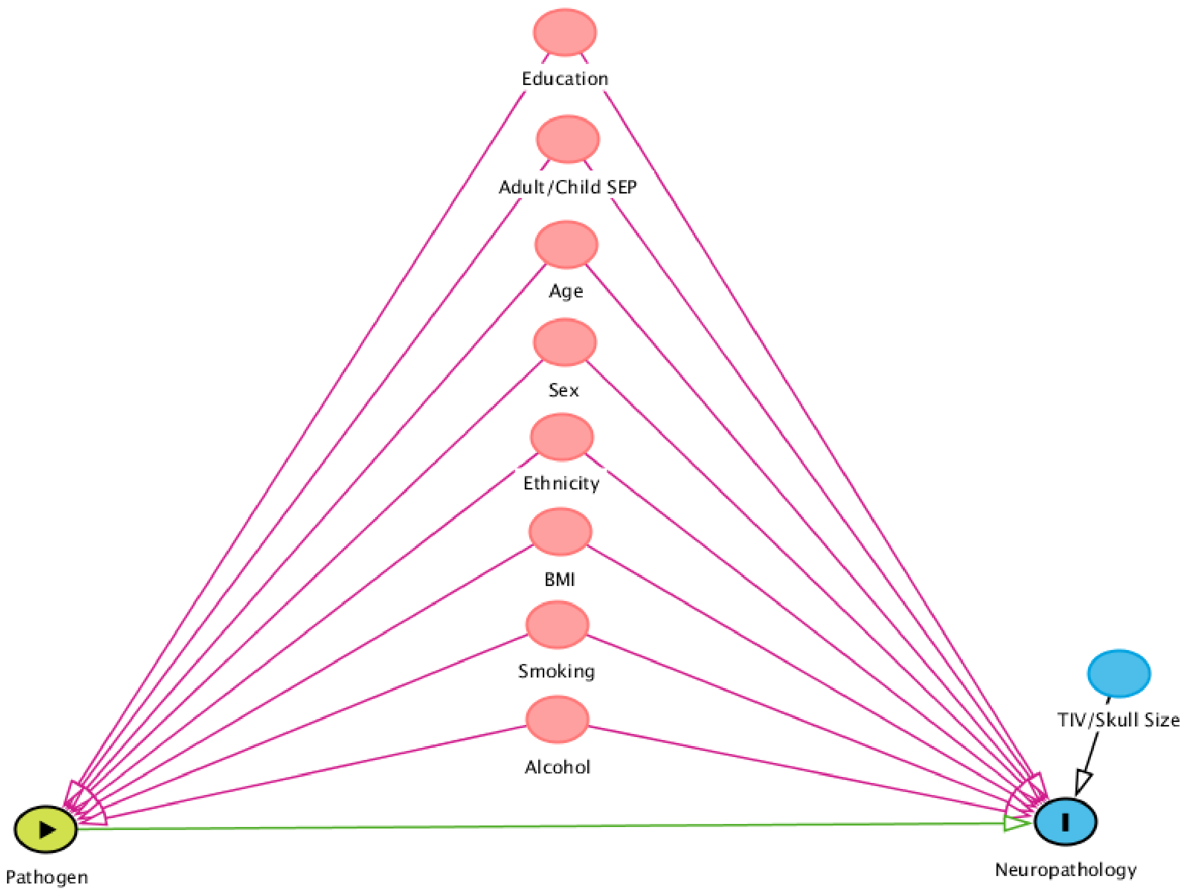

**Sensitivity analyses**

We considered diabetes, immunosuppressant medication, liver disease, chronic kidney disease, rheumatic heart disease, and autoimmune diseases as possible exclusion criteria in a sensitivity analyses. However, of these variables, only diabetes was available in all cohorts.
