## Supplementary Figure for "Common infections and neuroimaging markers of dementia in three UK cohort studies"

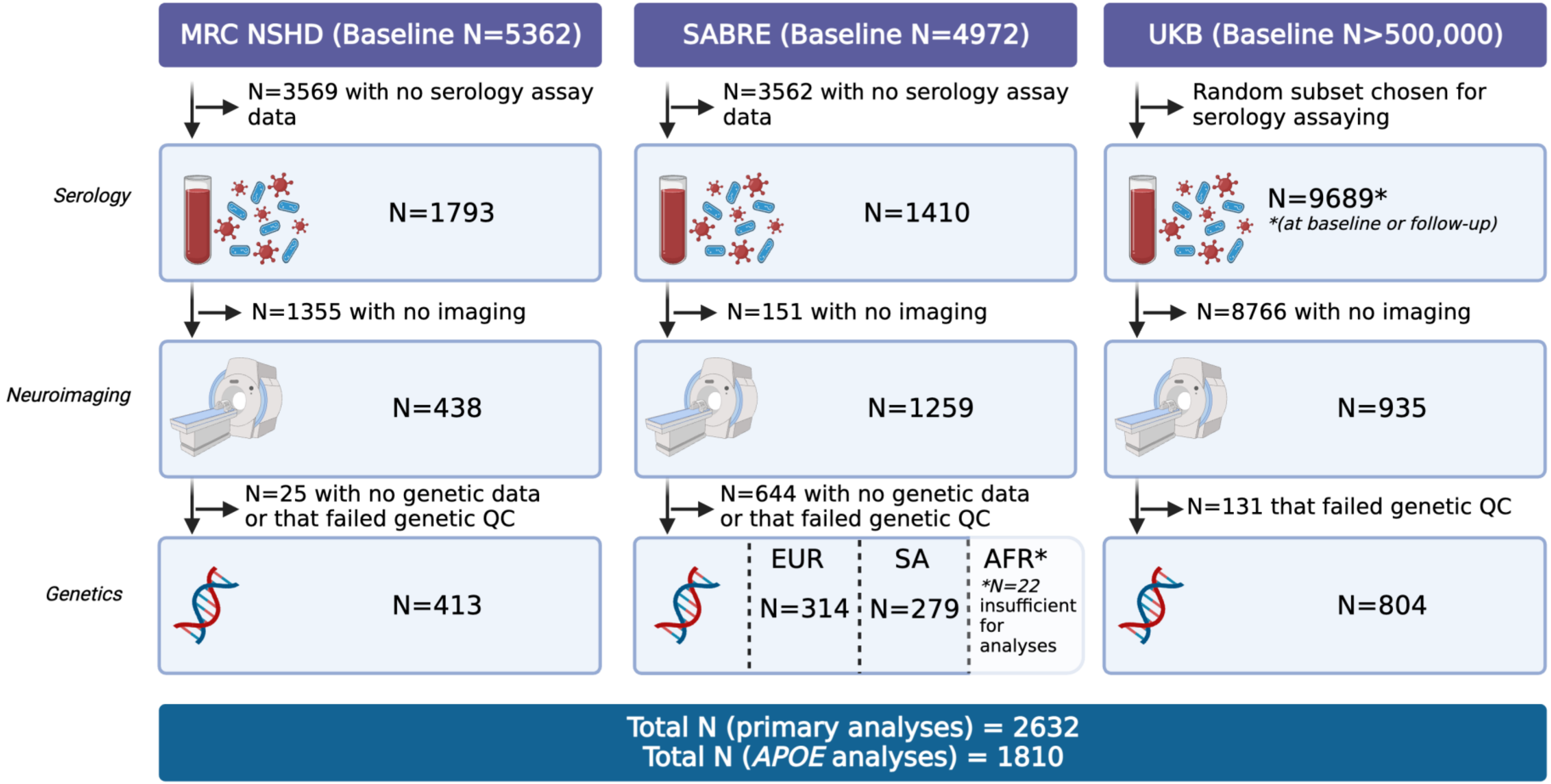


**Supplementary Figure 1. Overview of study participants.**
Abbreviations: AFR=African Caribbean, *APOE*=Apolipoprotein E genotype, EUR=European, MRC NSHD=Medical Research Council National Survey of Health and Development, SA=South Asian, SABRE=Southall and Brent Revisited, QC=quality control, UKB=UK Biobank.

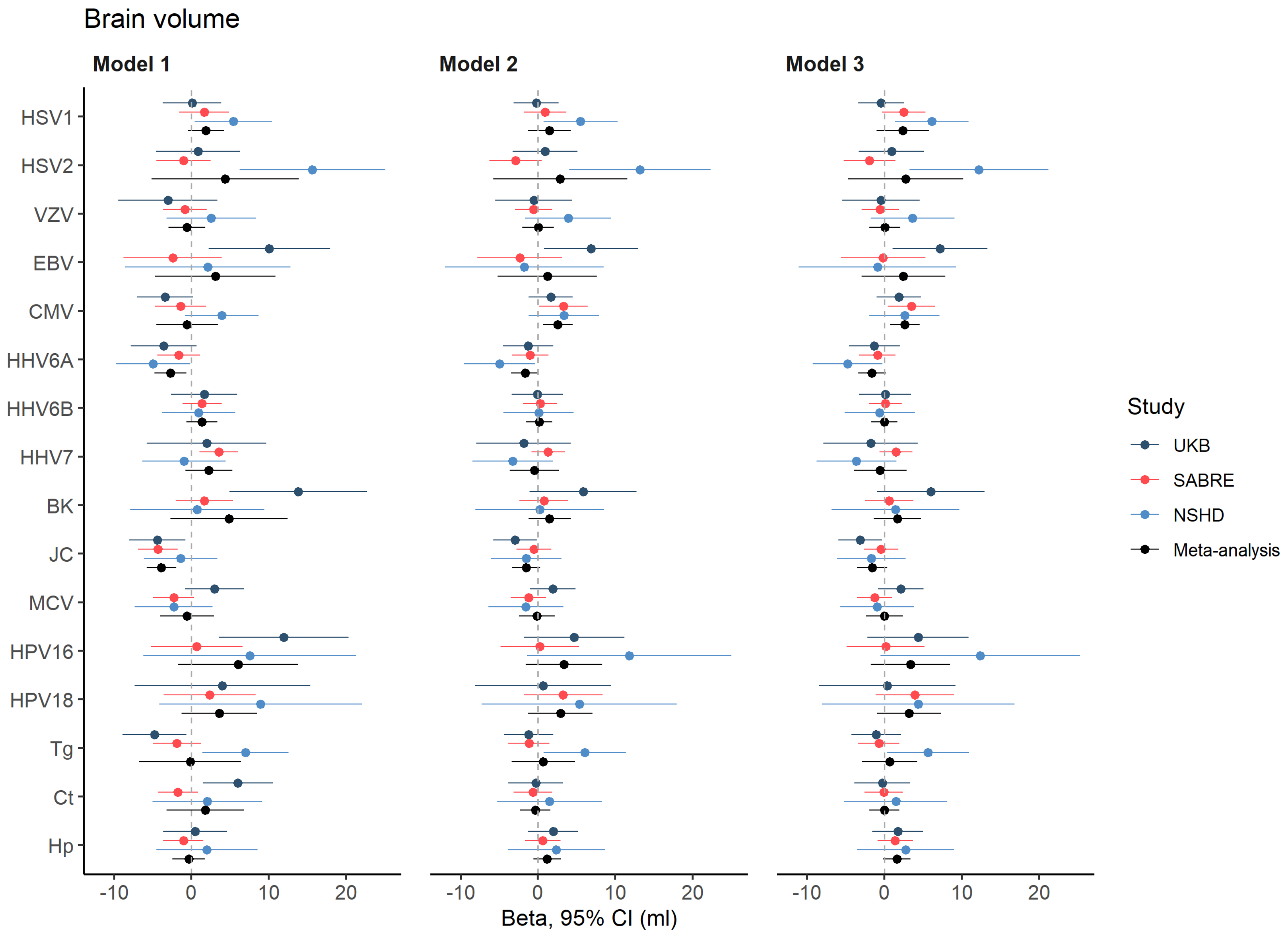


**Supplementary Figure 2. Forest plot indicating per-cohort and meta-analysed associations of pathogen serostatus with brain volume.** Model 1 included adjustments for total intracranial volume and other technical covariates; model 2 additionally for age, sex, and ethnicity; and model 3 additionally for BMI, smoking status, educational attainment, socioeconomic position, and alcohol intake. Studies are indicated by colour.

Abbreviations: BK=BK virus; CMV=Cytomegalovirus; Ct=*C.trachomatis,* EBV=Epstein-Barr virus; HHV=Human herpesvirus; Hp=*H.pylori;* HPV=Human papillomavirus; HSV=Herpes-simplex virus; JC=John Cunningham virus; MCV=Merkel Cell virus; NSHD=National Survey of Health and Development; SABRE=Southall and Brent Revisited; Tg=*T.gondii;* UKB=UK Biobank; VZV=Varicella zoster virus.

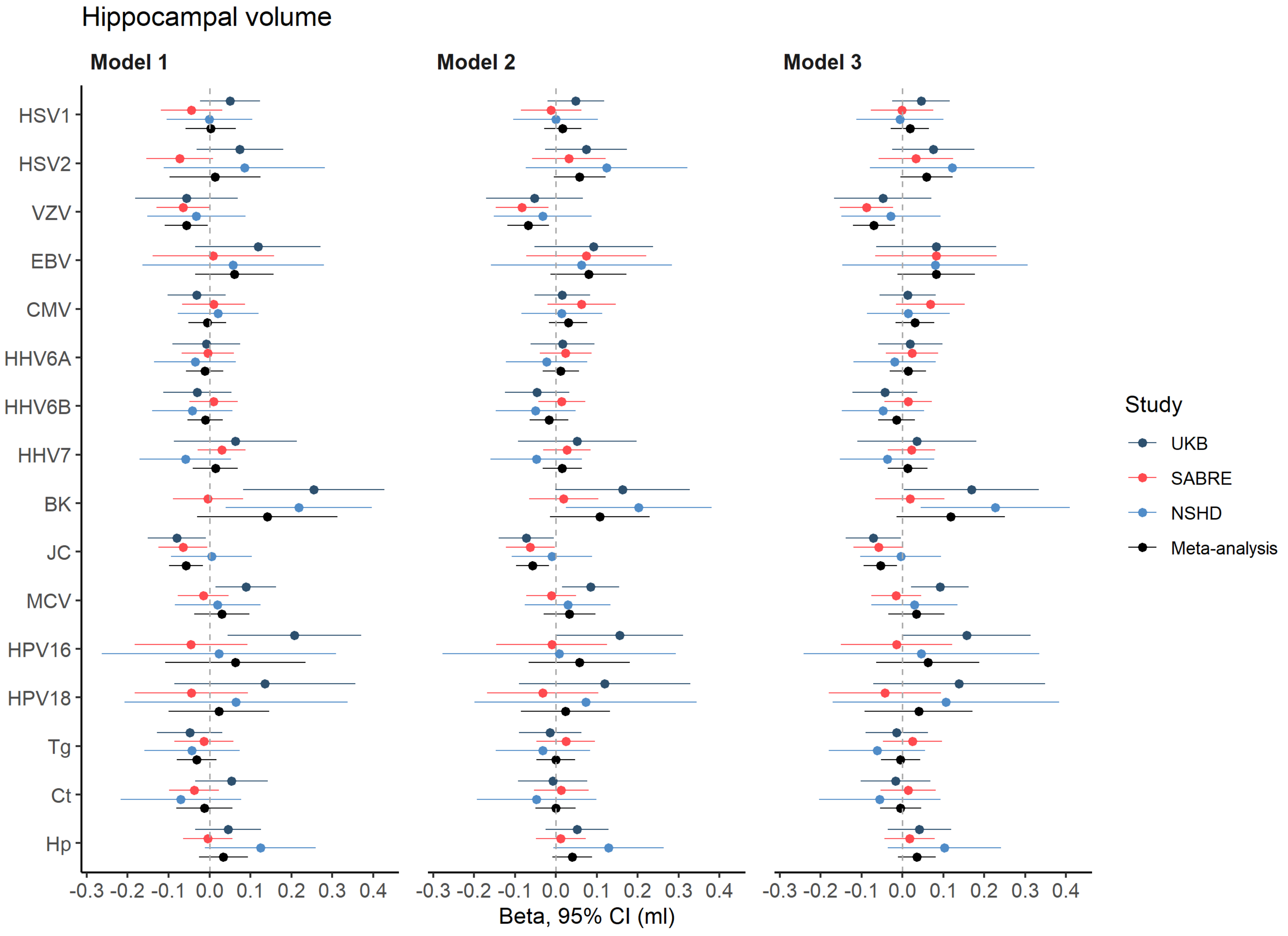


**Supplementary Figure 3. Forest plot indicating per-cohort and meta-analysed associations of pathogen serostatus with hippocampal volume.** Model 1 included adjustments for total intracranial volume and other technical covariates; model 2 additionally for age, sex, and ethnicity; and model 3 additionally for BMI, smoking status, educational attainment, socioeconomic position, and alcohol intake. Studies are indicated by colour.

Abbreviations: BK=BK virus; CMV=Cytomegalovirus; Ct=*C.trachomatis,* EBV=Epstein-Barr virus; HHV=Human herpesvirus; Hp=*H.pylori;* HPV=Human papillomavirus; HSV=Herpes-simplex virus; JC=John Cunningham virus; MCV=Merkel Cell virus; NSHD=National Survey of Health and Development; SABRE=Southall and Brent Revisited; Tg=*T.gondii;* UKB=UK Biobank; VZV=Varicella zoster virus.

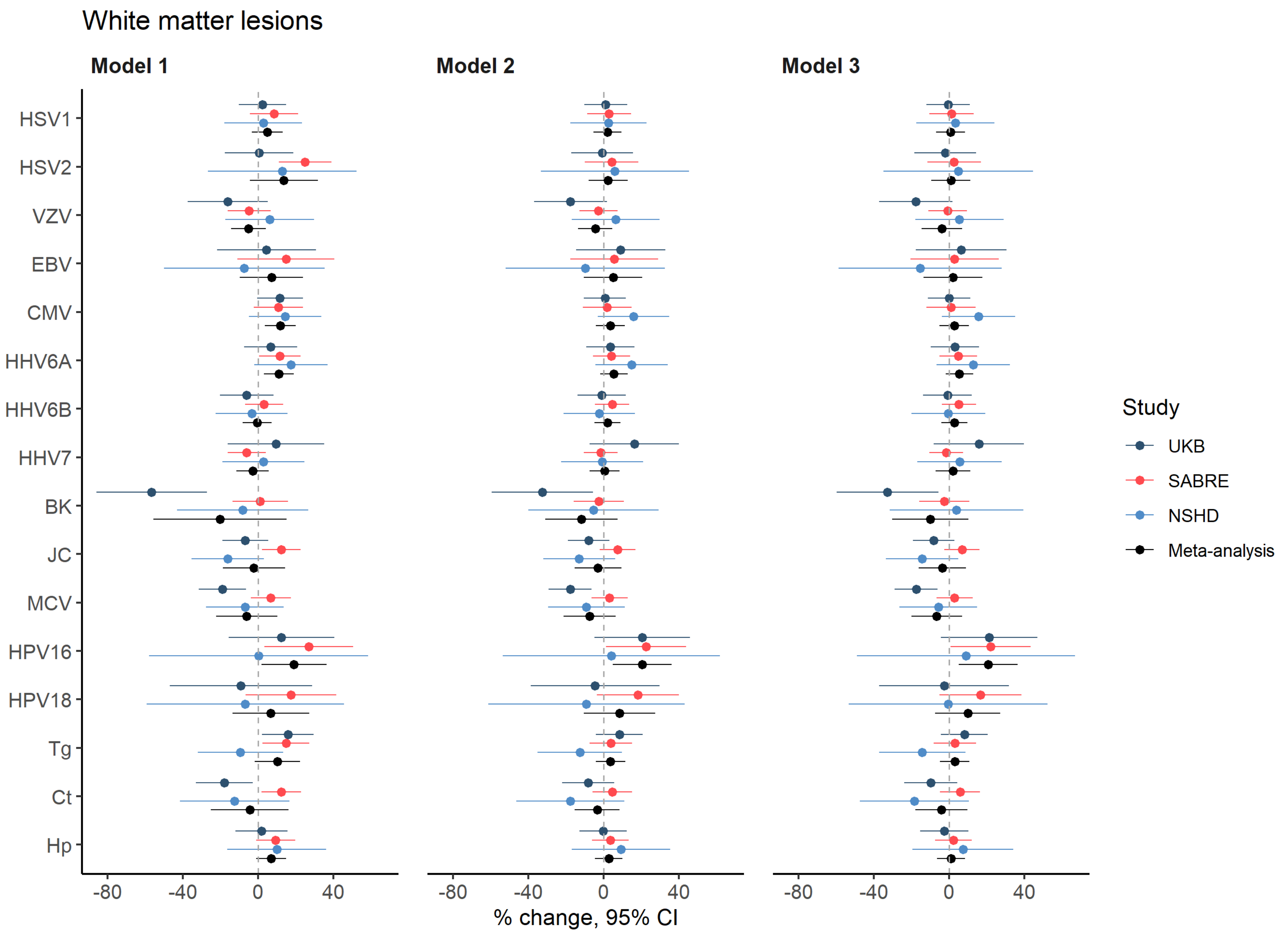


**Supplementary Figure 4. Forest plot indicating per-cohort and meta-analysed associations of pathogen serostatus with white matter lesions.** Model 1 included adjustments for total intracranial volume and other technical covariates; model 2 additionally for age, sex, and ethnicity; and model 3 additionally for BMI, smoking status, educational attainment, socioeconomic position, and alcohol intake. Studies are indicated by colour.

Abbreviations: BK=BK virus; CMV=Cytomegalovirus; Ct=*C.trachomatis,* EBV=Epstein-Barr virus; HHV=Human herpesvirus; Hp=*H.pylori;* HPV=Human papillomavirus; HSV=Herpes-simplex virus; JC=John Cunningham virus; MCV=Merkel Cell virus; NSHD=National Survey of Health and Development; SABRE=Southall and Brent Revisited; Tg=*T.gondii;* UKB=UK Biobank; VZV=Varicella zoster virus.

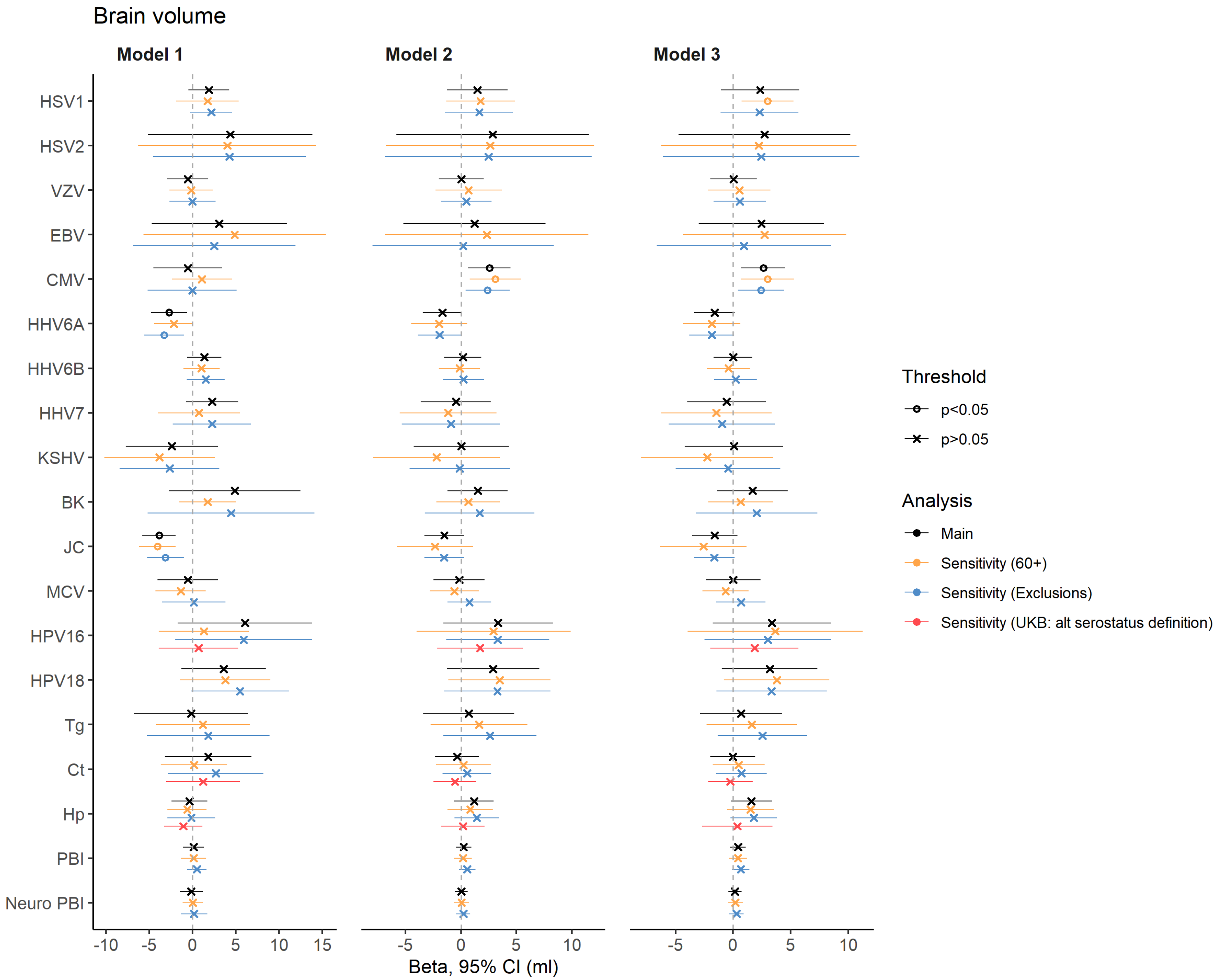


**Supplementary Figure 5. Forest plot indicating results from primary and sensitivity analyses of pathogen serostatus with brain volume.** Model 1 included adjustments for total intracranial volume and other technical covariates; model 2 additionally for age, sex, and ethnicity; and model 3 additionally for BMI, smoking status, educational attainment, socioeconomic position, and alcohol intake. Analyses are indicated by colour.

Abbreviations: BK=BK virus; CMV=Cytomegalovirus; Ct=*C.trachomatis,* EBV=Epstein-Barr virus; HHV=Human herpesvirus; Hp=*H.pylori;* HPV=Human papillomavirus; HSV=Herpes-simplex virus; JC=John Cunningham virus; MCV=Merkel Cell virus; PBI=pathogen burden index; Tg=*T.gondii;* UKB=UK Biobank;VZV=Varicella zoster virus.

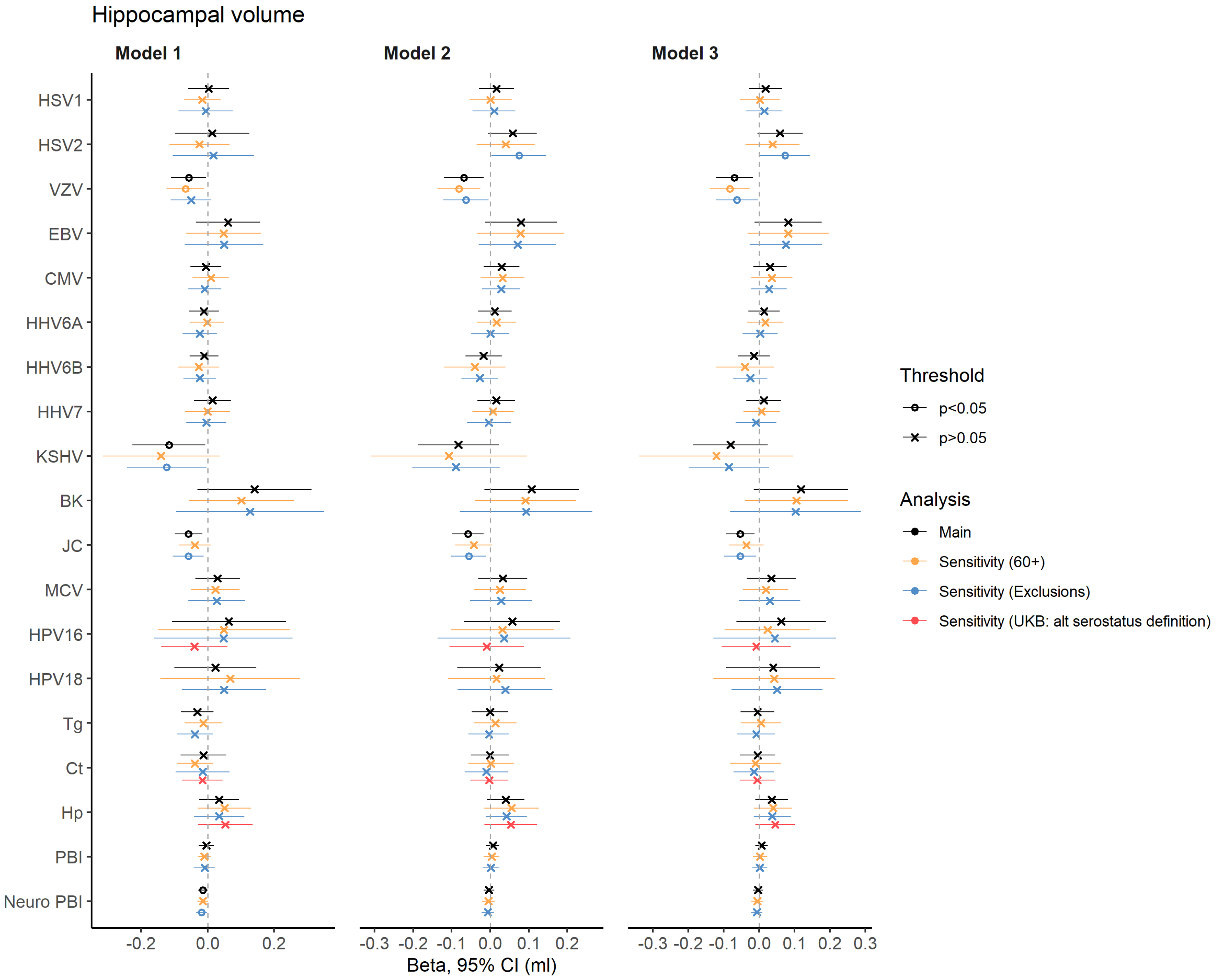


**Supplementary Figure 6. Forest plot indicating results from primary and sensitivity analyses of pathogen serostatus with hippocampal volume.** Model 1 included adjustments for total intracranial volume and other technical covariates; model 2 additionally for age, sex, and ethnicity; and model 3 additionally for BMI, smoking status, educational attainment, socioeconomic position, and alcohol intake. Analyses are indicated by colour.

Abbreviations: BK=BK virus; CMV=Cytomegalovirus; Ct=*C.trachomatis,* EBV=Epstein-Barr virus; HHV=Human herpesvirus; Hp=*H.pylori;* HPV=Human papillomavirus; HSV=Herpes-simplex virus; JC=John Cunningham virus; MCV=Merkel Cell virus; PBI=pathogen burden index; Tg=*T.gondii;* UKB=UK Biobank;VZV=Varicella zoster virus.

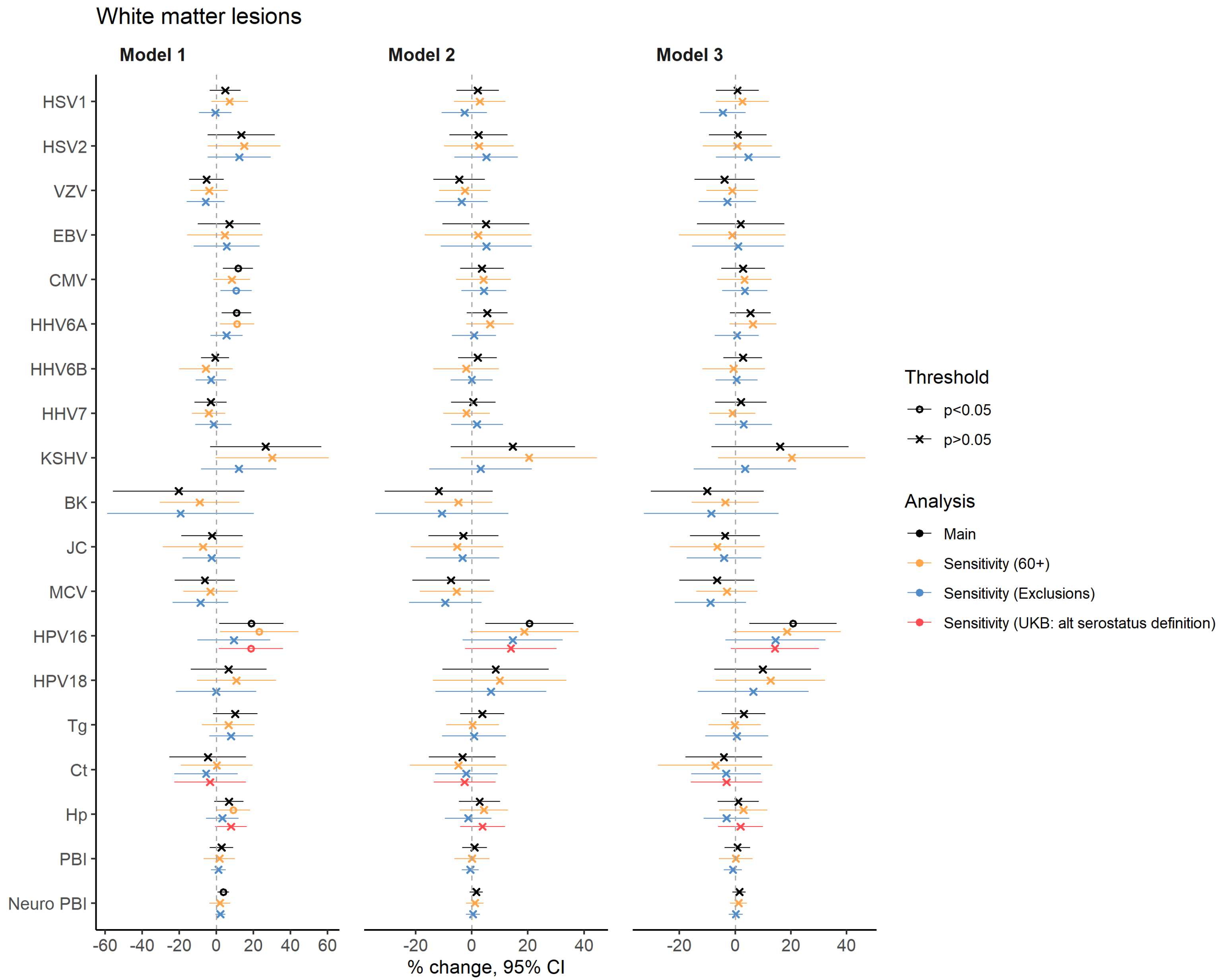


**Supplementary Figure 7. Forest plot indicating results from primary and sensitivity analyses of pathogen serostatus with white matter lesions.** Model 1 included adjustments for total intracranial volume and other technical covariates; model 2 additionally for age, sex, and ethnicity; and model 3 additionally for BMI, smoking status, educational attainment, socioeconomic position, and alcohol intake. Analyses are indicated by colour.

Abbreviations: BK=BK virus; CMV=Cytomegalovirus; Ct=*C.trachomatis,* EBV=Epstein-Barr virus; HHV=Human herpesvirus; Hp=*H.pylori;* HPV=Human papillomavirus; HSV=Herpes-simplex virus; JC=John Cunningham virus; MCV=Merkel Cell virus; PBI=pathogen burden index; Tg=*T.gondii;* UKB=UK Biobank;VZV=Varicella zoster virus.

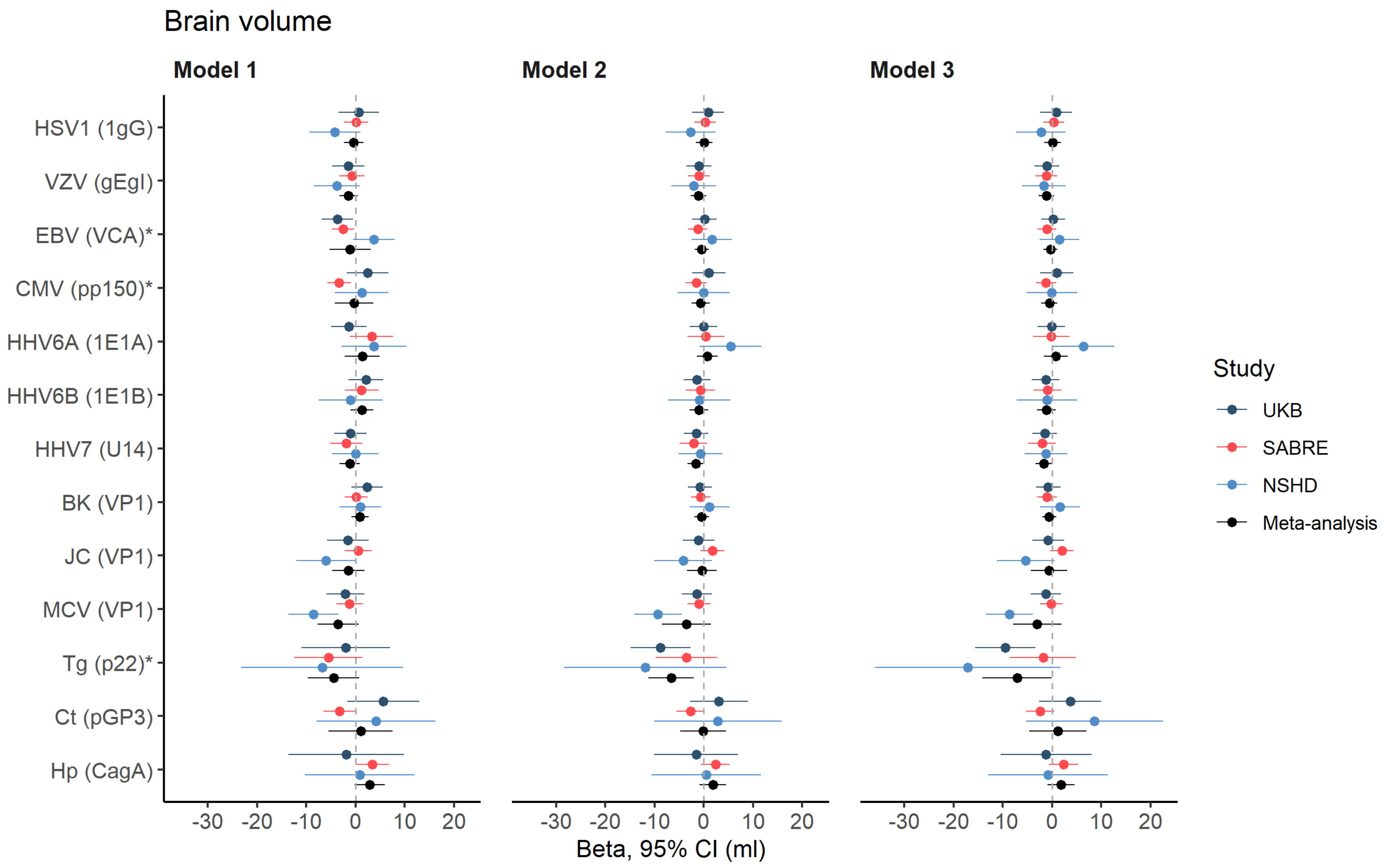


**Supplementary Figure 8. Forest plot indicating per-cohort and meta-analysed associations of seroreactivity tertiles with brain volume.** Model 1 included adjustments for total intracranial volume and other technical covariates; model 2 additionally for age, sex, and ethnicity; and model 3 additionally for BMI, smoking status, educational attainment, socioeconomic position, and alcohol intake. Pathogens and specific antigens (in brackets) are indicated on the y-axis. Studies are indicated by colour. HSV2 not shown due to large confidence intervals rendering plot uninterpretable, however these data are available on request.

Abbreviations: BK=BK virus; CMV=Cytomegalovirus; Ct=*C.trachomatis*, EBV=Epstein-Barr virus; HHV=Human herpesvirus; Hp=*H.pylori;* HPV=Human papillomavirus; HSV=Herpes-simplex virus; JC=John Cunningham virus; MCV=Merkel Cell virus; NSHD=National Survey of Health and Development; SABRE=Southall and Brent Revisited; Tg=*T.gondii*; UKB=UK Biobank; VZV=Varicella zoster virus.

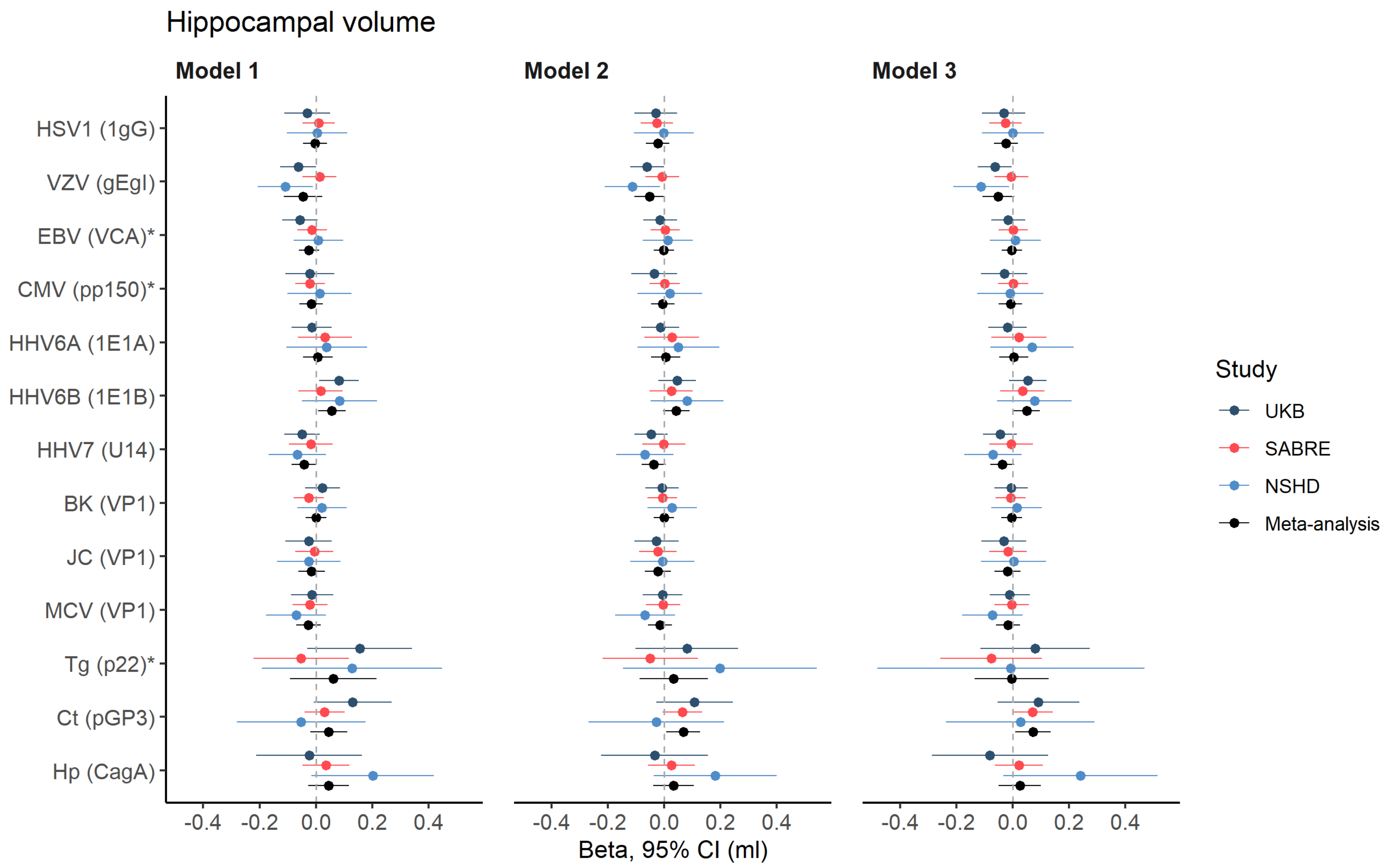


**Supplementary Figure 9. Forest plot indicating per-cohort and meta-analysed associations of seroreactivity tertiles with hippocampal volume.** Model 1 included adjustments for total intracranial volume and other technical covariates; model 2 additionally for age, sex, and ethnicity; and model 3 additionally for BMI, smoking status, educational attainment, socioeconomic position, and alcohol intake. Pathogens and specific antigens (in brackets) are indicated on the y-axis. Studies are indicated by colour. HSV2 not shown due to large confidence intervals rendering plot uninterpretable, however these data are available on request. Abbreviations: BK=BK virus; CMV=Cytomegalovirus; Ct=*C.trachomatis*, EBV=Epstein-Barr virus; HHV=Human herpesvirus; Hp=*H.pylori;* HPV=Human papillomavirus; HSV=Herpes-simplex virus; JC=John Cunningham virus; MCV=Merkel Cell virus; NSHD=National Survey of Health and Development; SABRE=Southall and Brent Revisited; Tg=*T.gondii*; UKB=UK Biobank; VZV=Varicella zoster virus.

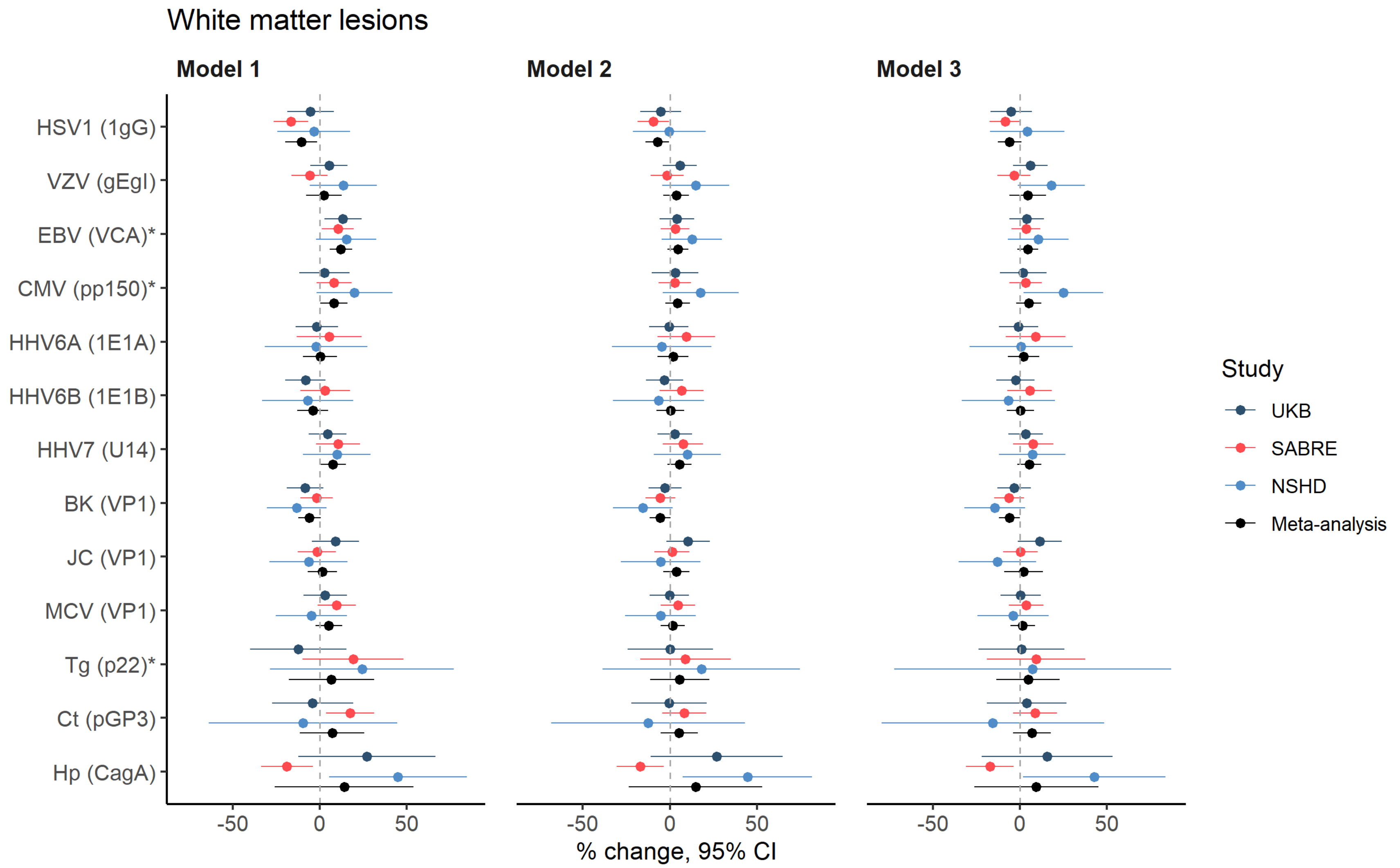


**Supplementary Figure 10. Forest plot indicating per-cohort and meta-analysed associations of seroreactivity tertiles with white matter lesions.** Model 1 included adjustments for total intracranial volume and other technical covariates; model 2 additionally for age, sex, and ethnicity; and model 3 additionally for BMI, smoking status, educational attainment, socioeconomic position, and alcohol intake. Pathogens and specific antigens (in brackets) are indicated on the y-axis. Studies are indicated by colour. HSV2 not shown due to large confidence intervals rendering plot uninterpretable, however these data are available on request. Abbreviations: BK=BK virus; CMV=Cytomegalovirus; Ct=*C.trachomatis*, EBV=Epstein-Barr virus; HHV=Human herpesvirus; Hp=*H.pylori;* HPV=Human papillomavirus; HSV=Herpes-simplex virus; JC=John Cunningham virus; MCV=Merkel Cell virus; NSHD=National Survey of Health and Development; SABRE=Southall and Brent Revisited; Tg=*T.gondii*; UKB=UK Biobank; VZV=Varicella zoster virus.

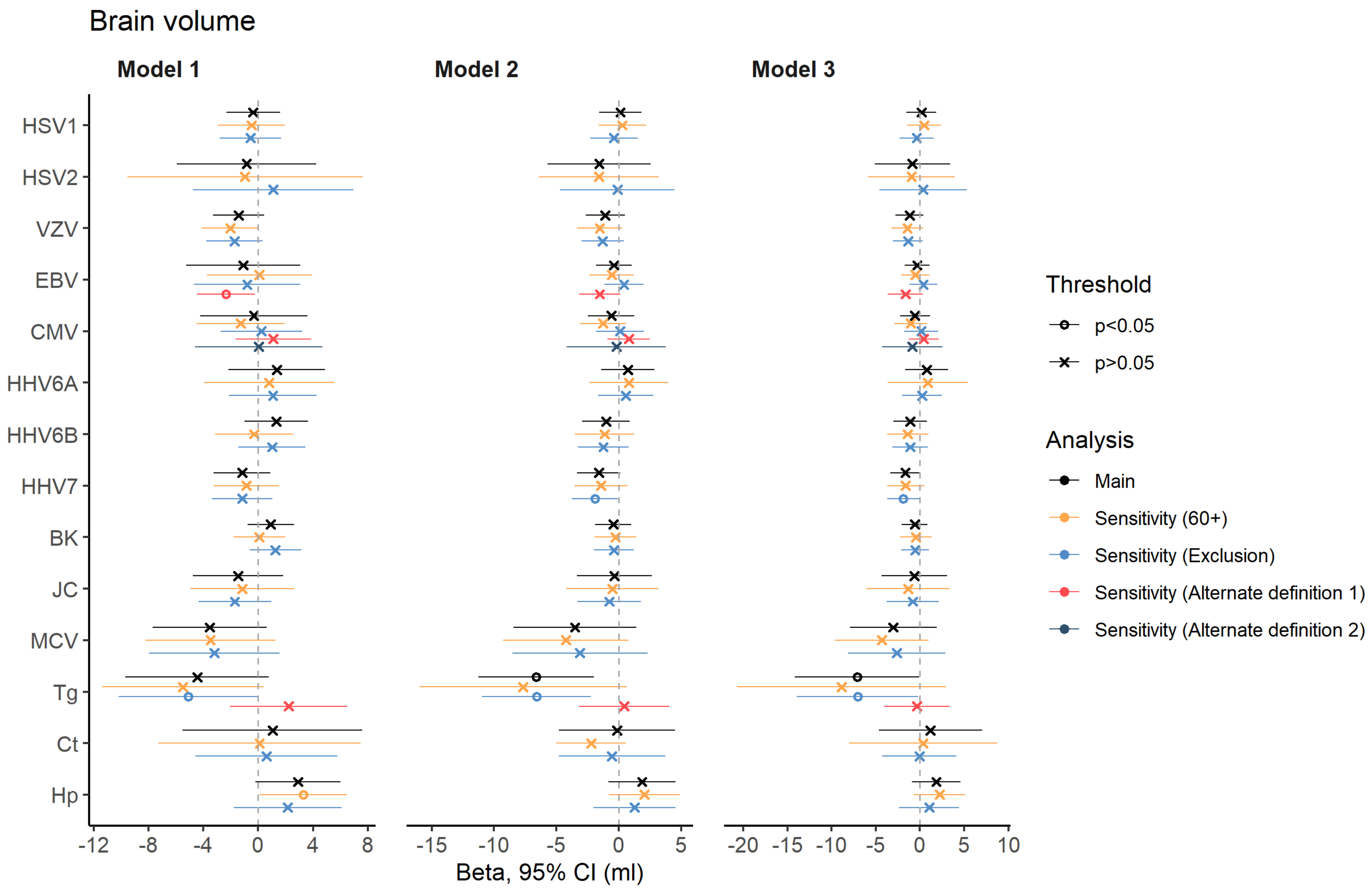


**Supplementary Figure 11. Forest plot indicating results from primary and sensitivity analyses of pathogen seroreactivity with brain volume.** Model 1 included adjustments for total intracranial volume and other technical covariates; model 2 additionally for age, sex, and ethnicity; and model 3 additionally for BMI, smoking status, educational attainment, socioeconomic position, and alcohol intake. Analyses are indicated by colour. Model 3 (sensitivity 60+) would not run in UK Biobank for model 3 due to insufficient degrees of freedom.

Abbreviations: BK=BK virus; CMV=Cytomegalovirus; Ct=Chlamydia trachomatis, EBV=Epstein-Barr virus; HHV=Human herpesvirus; Hp=Helicobacter pylori; HPV=Human papillomavirus; HSV=Herpes-simplex virus; JC=John Cunningham virus; MCV=Merkel Cell virus; Tg=Toxoplasma gondii; UKB=UK Biobank; VZV=Varicella zoster virus.

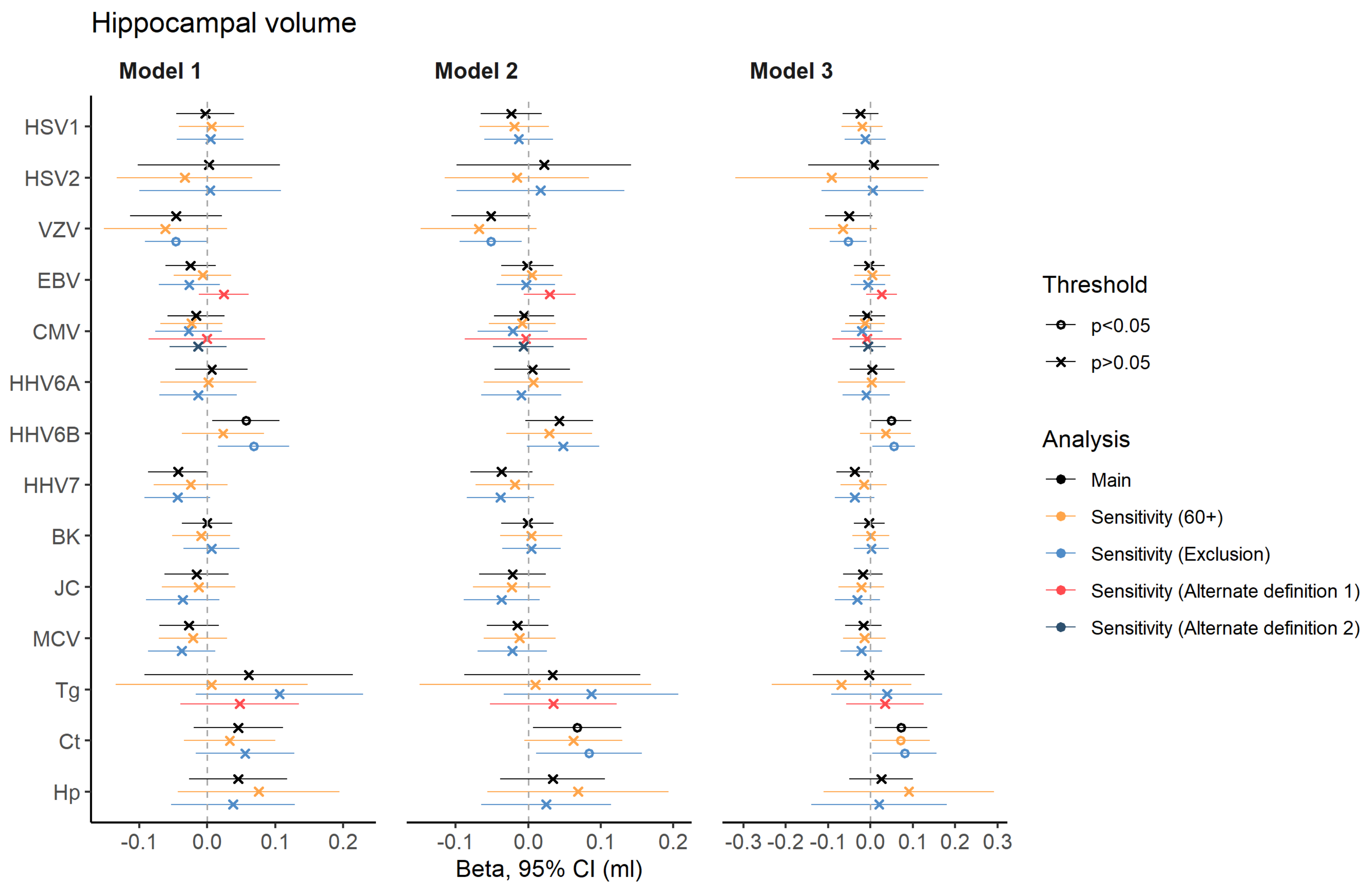


**Supplementary Figure 12. Forest plot indicating results from primary and sensitivity analyses of pathogen seroreactivity with hippocampal volume.** Model 1 included adjustments for total intracranial volume and other technical covariates; model 2 additionally for age, sex, and ethnicity; and model 3 additionally for BMI, smoking status, educational attainment, socioeconomic position, and alcohol intake. Analyses are indicated by colour. Model 3 (sensitivity 60+) would not run in UK Biobank for model 3 due to insufficient degrees of freedom.

Abbreviations: BK=BK virus; CMV=Cytomegalovirus; Ct=Chlamydia trachomatis, EBV=Epstein-Barr virus; HHV=Human herpesvirus; Hp=Helicobacter pylori; HPV=Human papillomavirus; HSV=Herpes-simplex virus; JC=John Cunningham virus; MCV=Merkel Cell virus; Tg=Toxoplasma gondii; UKB=UK Biobank; VZV=Varicella zoster virus.

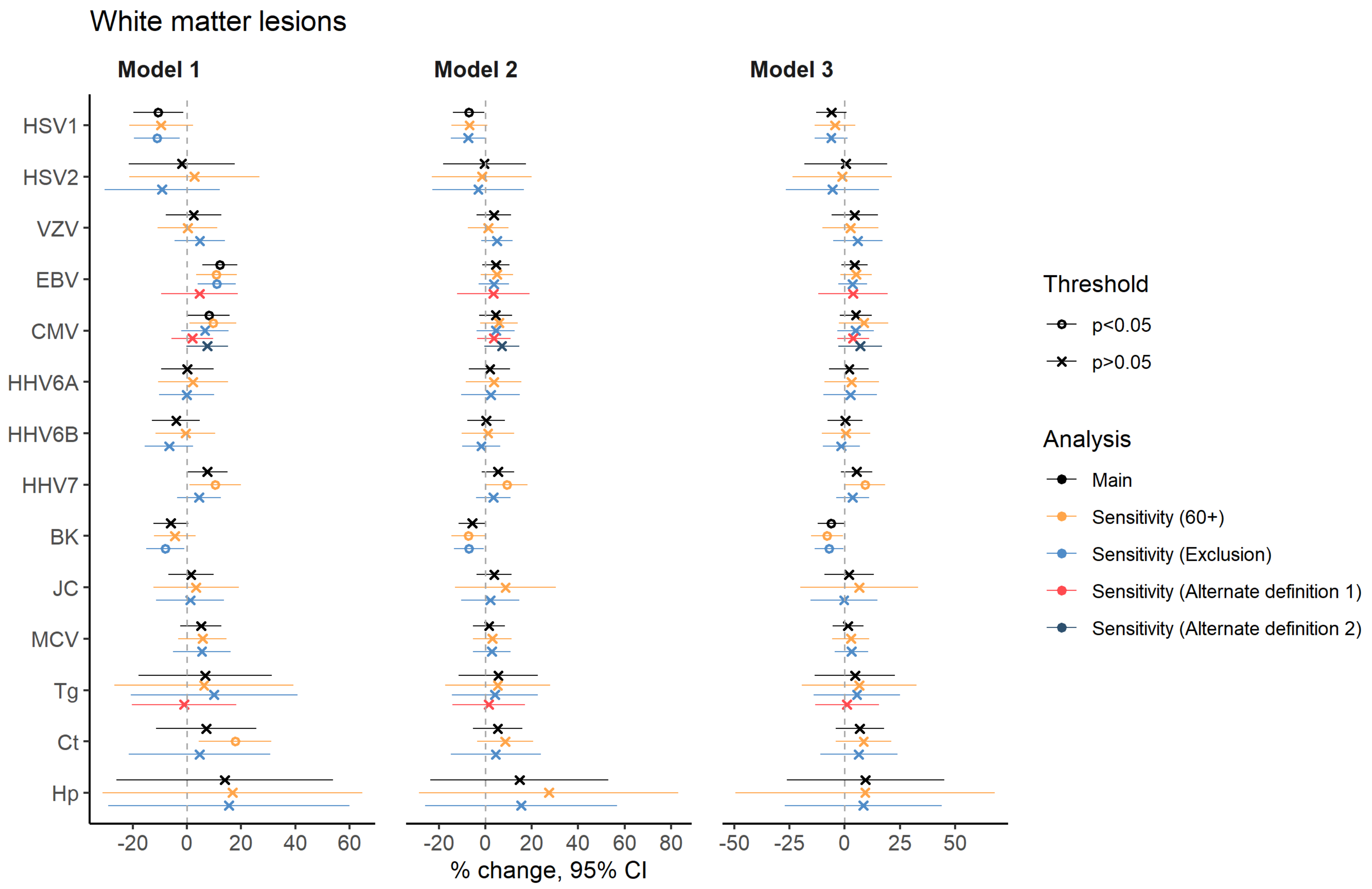


**Supplementary Figure 13. Forest plot indicating results from primary and sensitivity analyses of pathogen seroreactivity with white matter lesions.** Model 1 included adjustments for total intracranial volume and other technical covariates; model 2 additionally for age, sex, and ethnicity; and model 3 additionally for BMI, smoking status, educational attainment, socioeconomic position, and alcohol intake. Analyses are indicated by colour. Model 3 (sensitivity 60+) would not run in UK Biobank for model 3 due to insufficient degrees of freedom.

Abbreviations: BK=BK virus; CMV=Cytomegalovirus; Ct=Chlamydia trachomatis, EBV=Epstein-Barr virus; HHV=Human herpesvirus; Hp=Helicobacter pylori; HPV=Human papillomavirus; HSV=Herpes-simplex virus; JC=John Cunningham virus; MCV=Merkel Cell virus; Tg=Toxoplasma gondii; UKB=UK Biobank; VZV=Varicella zoster virus.
